# Pay-as-you-go LPG supports sustainable clean cooking in Kenyan informal urban settlement, including during a period of COVID-19 lockdown

**DOI:** 10.1101/2020.11.20.20235978

**Authors:** Matthew Shupler, Mark O’Keefe, Elisa Puzzolo, Emily Nix, Rachel Anderson de Cuevas, James Mwitari, Arthur Gohole, Edna Sang, Iva Čukić, Diana Menya, Daniel Pope

## Abstract

Approximately 2.8 billion people rely on polluting cooking fuels (e.g. wood, kerosene), exposing them to health-damaging household air pollution. A key access barrier to clean cooking fuels (e.g. liquefied petroleum gas (LPG)) is affordability. By enabling households to pay in small increments, pay-as-you-go (PAYG) LPG could help promote clean cooking, and support continued LPG use through periods of economic downturn. We investigate the ability of PAYG LPG to sustain access to clean cooking from January 2018-June 2020, including during COVID-19 lockdown (March-June 2020) in an informal settlement in Nairobi, Kenya. We utilize novel PAYG LPG smart meter data to document cooking/spending patterns from 426 PAYG LPG customers and semi-structured interviews among a subset of seven households. Objective cooking pattern comparisons are made to those cooking with full 6kg cylinder LPG and polluting fuel users from 23 households in peri-urban Eldoret in western Kenya, using stove monitoring data. Customers’ average PAYG LPG consumption was 0.97 kg/capita/month (11.6 kg/capita/year) prior to COVID-19 lockdown. Despite adverse economic impacts of the lockdown, 95% of households continued using PAYG LPG, and consumption increased to 1.22 kg/capita/month (March-June 2020). Daily cooking events using PAYG LPG increased by 60% (1.07 events/day (pre-lockdown) to 1.72 events/day (lockdown)). In contrast, among seven households purchasing full 6kg cylinder LPG in Eldoret, average days/month using LPG declined by 75% (17 to four days) during COVID-19 lockdown. Median PAYG LPG payment frequency doubled (from every 8 days to every 4 days) during lockdown, while average payment amount was nearly halved (336 Kenyan Shillings (KSh)/US$3.08 to 179 KSh/US$1.64).

Interviewed customers reported numerous benefits of PAYG LPG beyond fuel affordability, including safety, time savings, cylinder delivery and user-friendliness. PAYG LPG helped sustain clean cooking during COVID-19 lockdown, possibly averting increases in polluting cooking fuel use and associated household air pollution exposures.

**Graphical Abstract:** 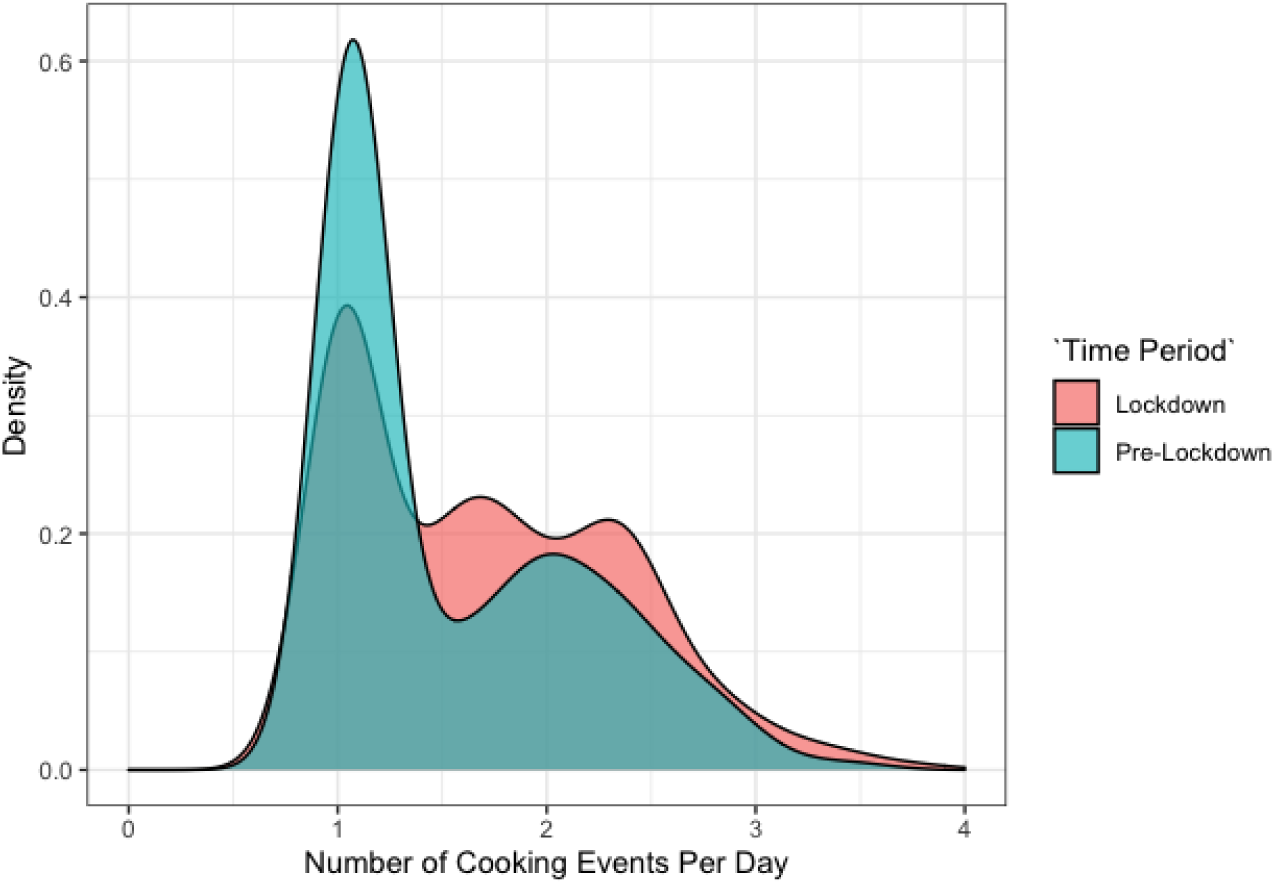

**Highlights:** First study to report long-term cooking/spending patterns using PAYG LPG smart meter data

95% of 301 active PAYG LPG customers in February 2020 continued to use the fuel during COVID-19 lockdown

Daily cooking events using PAYG LPG increased by 60% during COVID-19 lockdown

Median payment frequency increased 50%, while single payment amount decreased 50% during lockdown

Three-quarters of PAYG LPG households in this study were first time LPG users

## 1. Introduction

An estimated 2.8 billion people rely on polluting fuels, including solid fuels (e.g. wood, charcoal) and kerosene, for their household energy needs.^1^ Unsafe levels of fine particulate matter (PM2.5) generated from combustion of polluting fuels is an established risk factor for several infectious and non-communicable respiratory and cardiovascular diseases.^2–6^ Further, residential biomass combustion generates 25% of global emissions of black carbon (BC),^7^ the dark component of particulate matter and a short-lived pollutant that has strong visible light absorption properties.^7–9^ BC is estimated to have the second largest radiative forcing^8,10^ following only CO_2_ ^7,10^

Clean burning liquefied petroleum gas (LPG), although a fossil fuel, emits low levels of BC and minimal PM2.5 concentrations, typically meeting WHO Indoor Air Quality Guideline levels for health. LPG can therefore have a neutral or ‘cooling’ effect on climate by reducing emissions of BC when it replaces use of biomass fuels for cooking in households.^13–15^ Replacing biomass cooking fuels with LPG also reduces deforestation and associated emissions of CO2,^16–18^ and alleviates time poverty through decreased cooking time.^19^ LPG is currently used for cooking (exclusively or alongside polluting fuels^20,21^) by >2.5 billion people worldwide,^22^ especially in Latin America^23^ and with rapid expansion in India^24^ and Indonesia.^25^ In Sub-Saharan Africa (SSA), however, 85% of the population relies on polluting cooking fuels, twice the global average (40%).^26^

### 1.1 Barriers to using liquefied petroleum gas for clean cooking

Polluting cooking fuels are commonly used in SSA due to their availability and low cost, or the ability to gather biomass for free from local forested areas, particularly in East Africa.^17,27^ While LPG costs are frequently lower relative to purchased biomass fuels on a per kilogram basis,^28^ an unreliable supply of LPG and the financial outlay required to pay upfront for large units of gas (e.g. 6kg-15kg cylinders) under the standard branded cylinder recirculation model (BCRM)^29^ is prohibitive for many.^30^ Among those that can afford the initial cost of the full LPG cylinder, insecure incomes and precarious financial circumstances may prevent some households from seeking full cylinder LPG refills, leading to its unsustainable usage as a primary cooking fuel.^24^ Thus, year-round affordability of LPG must be achieved to facilitate universal access to affordable, reliable, sustainable and modern household energy (Sustainable Development Goal (SDG) 7), particularly when resource poor households are confronted with economic downturns, such as those due to COVID-19 control measures.^15,31^ A recent longitudinal study showed that 95% (n=183) of households in an informal settlement in Nairobi, Kenya experienced reductions in household income during COVID-19 lockdown, and 27% of full cylinder LPG users in the community reverted to kerosene (14%) or wood (13%) for cooking.^32^

### 1.2 Pay-as-you-go liquefied petroleum gas

Pay-as-you-go (PAYG) is a consumer finance mechanism that can potentially relieve the financial barrier to sustained clean energy access by allowing consumers to purchase LPG credits in small increments (via mobile banking). Pay-as-you-go smart meter technology has already been used to provide resource-poor households in SSA with affordable access to technologies for electricity and water/sanitation.^33–35^ From a social perspective, similarities of PAYG with the concept of paying for an energy service may help explain its success.^34^ Further, consumers/retailers can track LPG consumption in real-time to ensure timely home deliveries of refills, which eliminates the need to travel to retail locations and minimizes safety risks from illegal cylinder refills. Further, as income generated in the informal or agricultural sector can vary seasonally and influence cooking fuel use,^24^ PAYG LPG may offer families the payment flexibility needed to maintain clean cooking during periods of reduced household income. As several commercial companies have penetrated the clean cooking fuel market in East Africa,^28^ PAYG LPG smart meter technology may be a scalable consumer finance mechanism in SSA and one of several tools available for helping countries (e.g. Kenya, Ghana, Cameroon) achieve their ambitious targets for rapid market expansion of LPG by 2030.^36^

### 1.3 Study aims

Smart meter data was analyzed from 426 customers of PayGo Energy in Nairobi, Kenya, one of the first PAYG LPG company in Africa, in order to achieve two research goals: (1) to characterize PAYG LPG stove use and payment patterns to assess the ability of PAYG LPG to sustain clean cooking in SSA and (2) to quantify the effects of the COVID-19 community lockdown on PAYG LPG cooking behaviors. This study is one of the first, to the authors’ knowledge, to use PAYG LPG smart meter data to assess consumer spending and cooking patterns.

## 2. Methods

### 2.1 Study setting and timeline

PayGo Energy was founded in 2016 to offer pay-as-you-go (PAYG) LPG with stainless steel double burner cookstoves (supplied by *Real Flame*, based in India) to residential and commercial (small corner shops selling cooked foods) customers in Mukuru kwa Reuben informal settlement in Nairobi, Kenya. Mukuru kwa Reuben has over 0.5 million residents, and is one of many informal settlements in the city. In March 2017, 50 PayGo Energy customers in Mukuru kwa Reuben were initially enrolled without any metering technology (see Figure S1 in Supplemental Information for timeline). In August 2017, new customers registered with the company and were supplied with metered LPG via a diaphragm (“Goldcard”) meter, capable of measuring gas consumption to the nearest 0.2 kilograms. Beginning in 2019, the Goldcard meters were gradually swapped to more highly calibrated ‘cylinder smart meters’ that measured customer LPG fuel usage to the nearest 0.001 kilograms.

### 2.2 Statistical analysis of pay-as-you-go LPG smart meter data

While PayGo Energy customer LPG consumption and expenditure data was available from August 2017 through June 2020, data from 2017 (before the Goldcard meters were installed) were excluded (n=39; 8% of total sample) due to inaccurate information on LPG consumption. Customer payment and consumption data from Goldcard and smart meters in 2018-2020 was combined to increase the power of the analysis (sensitivity analyses examined data separately for each type of meter to assess robustness of results). In all analyses, separate consumption readings from the same PAYG LPG stove within the same hour were counted as a single cooking event; for example if three 0.01 kg usage events occurred within the same hour, this was considered as a single 0.03 kg cooking event.

In Kenya, a nationwide mandatory lockdown was enforced beginning on March 25, 2020. Two weeks later, on April 7, 2020, a dusk-to-dawn curfew (7 pm-5 am) was implemented. PayGo Energy consumer fuel usage and payment data was therefore separated into pre-COVID-19 lockdown months (January 2018-February 2020), and months during community lockdown (March -June 2020) to examine potential effects of the nationwide lockdown (and associated impacts on household incomes and food insecurity) on patterns of LPG usage. At time of registration for PayGo Energy equipment, new customers completed a baseline questionnaire on demographics and socioeconomic status (SES), with different information collected from enrolled customers depending on the year; the association of various consumer demographics and SES characteristics with PAYG LPG consumption and expenditures was examined.

### 2.3 Customer interviews

On August 18, 2020 (after the analysis period of smart meter data included in this study), a sample of seven customers (six residential and one commercial) of PayGo Energy were interviewed in-house to assess how the pandemic has affected their livelihoods; the interviewed customers were purposively selected from different areas of the study setting. The 10-minute telephonic, semi-structured interviews encompassed the following questions: (1) how has COVID-19 impacted you and your community? (2) have you changed your spending on LPG during COVID-19 and why? (3) do you cook with other fuels in addition to PAYG LPG? and (4) have your cooking fuel choices changed during COVID-19? The interviews were conducted in Kiswahili by two local staff members employed by PayGo Energy (customer care associate and communications associate). After translation into English by native speakers of Kiswahili, key findings from the interviews were independently identified following the process of thematic analysis^37^ in computer-assisted qualitative data analysis software (ATLAS.ti Scientific Software Development GmbH). The results of the interviews were integrated with quantitative data to provide context for PAYG LPG cooking patterns before and during COVID-19 lockdown.

### 2.4 Stove use monitoring data in Eldoret, Kenya

In a separate study, stove use monitoring (SUM) data was collected from a sample of 23 households in Eldoret, a peri-urban town five hours drive away from Nairobi in Western Kenya. Stove temperature data was recorded every five minutes using temperature sensors (Geocene Dots) placed a standardized distance of 15 centimetres away from the centre of the flame on wood, charcoal and LPG stoves. Geocene Dots remained on the primary stoves used in these households for several months (November 2019-June 2020) prior to COVID-19 and during the community lockdown period. Temperature data was dichotomized into ‘stove on’ or ‘stove off’ based on a machine learning algorithm developed by Geocene.^38^ The SUM data analysis provided an objective comparison of cooking patterns from full 6kg cylinder LPG users relative to the PAYG LPG users in Kenya.

### 2.5 Data sharing

The anonymized PayGo Energy customer database was stored on a secure, cloud-based server hosted on the Google Cloud Platform and shared securely with University of Liverpool using

DatAnywhere. The interview recordings were securely shared with Liverpool using SharePoint. All statistical analyses were completed in R version 3.5.1.^39^ Ethical approval for this study was obtained from the University of Liverpool Central Ethics Committee, United Kingdom.

## 3. Results

### 3.1 Study population

Over 100,000 (n=135,353) days of LPG customer data on PAYG LPG usage from 426 PayGo Energy customers (415 residential (97%) and 11 commercial (3%)) from January 2018 – June 2020 were analysed. Customers were primarily recruited in two separate ‘waves’ in 2017 and 2019. Thus, the distribution of months of smart meter data available per customer was bimodal with peaks at 8 months and 34 months (range: 1-41 months) (Table S6). There were 288 (68%) active customers as of June 30, 2020. Of the 136 (133 residential, 3 commercial) PayGo Energy account deactivations, 95% (n=130) occurred in 2018 or 2019 - the most common reasons being moving away from the community (n=41; 32%) or tampered/stolen equipment (n=34; 26%) (Table S7). Only 13 customers deactivated their account (n=6) or had no recorded PAYG LPG usage during the first three months (April-June 2020) of the COVID-19 lockdown (n=7). Thus, 95% of PayGo Energy customers that were active in March 2020 continued using PAYG LPG during the lockdown.

Half (49%; n=124) the PayGo Energy residential customers lived in households comprising one multi-purpose room (Table 1). Nearly half (44%; n=110) of female heads of household had not completed more than primary level education, compared with 28% (n=61) of male heads of household. A female was the main cook in four in five households (83%; n=211), yet the primary decision maker for choice of cooking fuel in only 59% (n=153) of households. This is, however, significantly higher (p=0.05) than the proportion of female cooking fuel decision makers among households in the informal urban settlement not using PAYG LPG (47%, n=49) (Table S3).

**Table 1.**
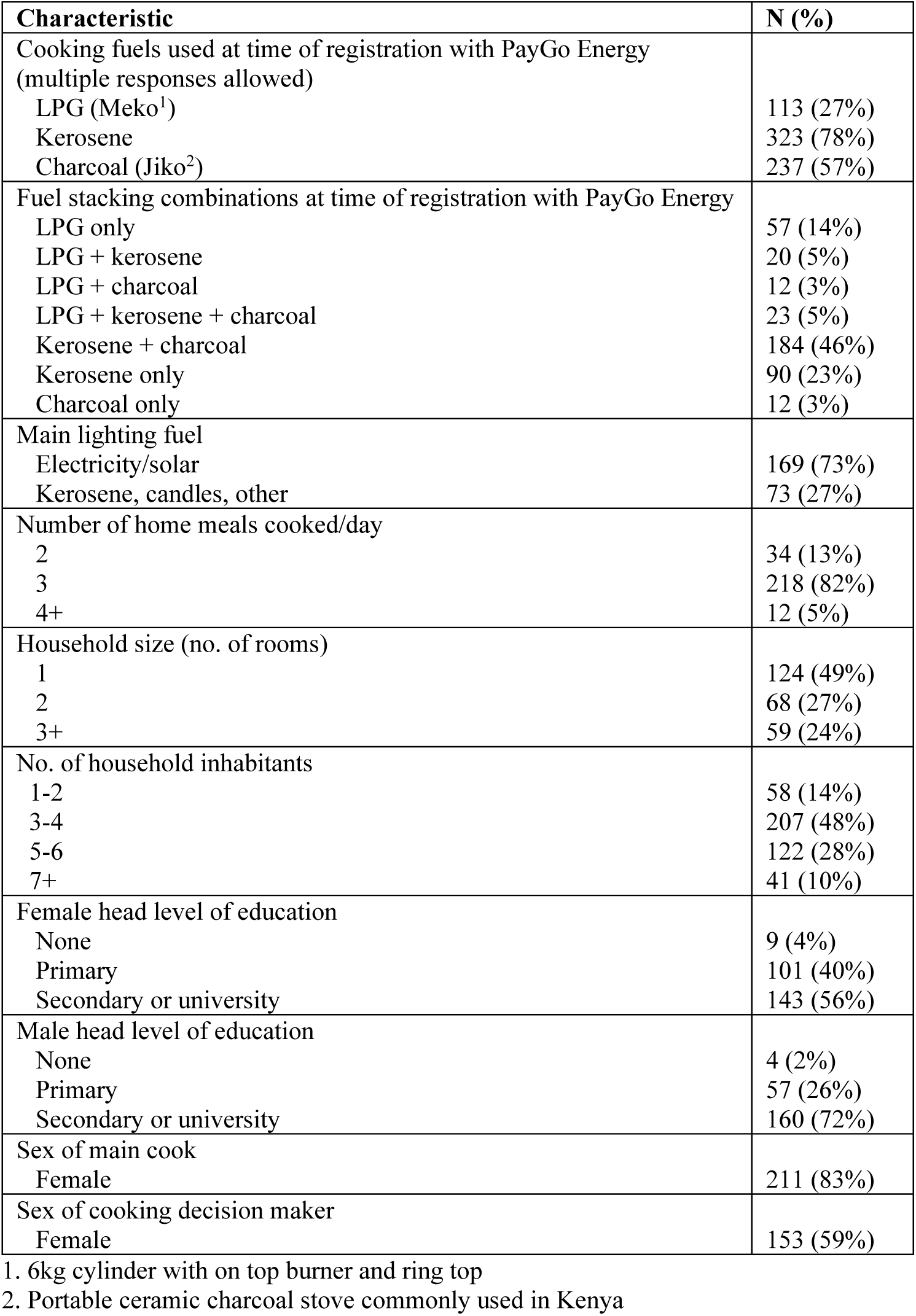
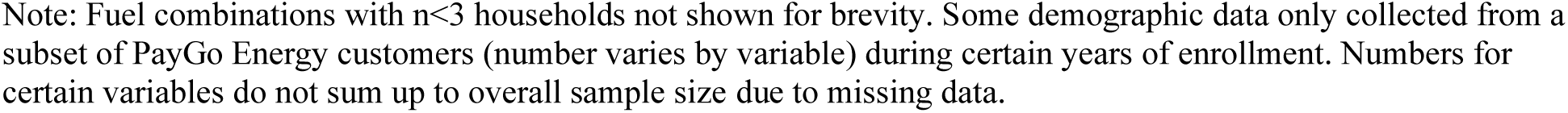
Demographics and cooking preferences at the time of registration for PAYG LPG among residential customers (all data collected before COVID-19 lockdown) (N=415) Note: Fuel combinations with n<3 households not shown for brevity. Some demographic data only collected from a subset of PayGo Energy customers (number varies by variable) during certain years of enrollment. Numbers for certain variables do not sum up to overall sample size due to missing data.

| Characteristic | N (%) |
| --- | --- |
| Cooking fuels used at time of registration with PayGo Energy<br>(multiple responses allowed) |  |
| LPG (Meko<sup>1</sup>) | 113 (27%) |
| Kerosene | 323 (78%) |
| Charcoal (Jiko<sup>2</sup>) | 237 (57%) |
| Fuel stacking combinations at time of registration with PayGo Energy |  |
| LPG only | 57 (14%) |
| LPG + kerosene | 20 (5%) |
| LPG + charcoal | 12 (3%) |
| LPG + kerosene + charcoal | 23 (5%) |
| Kerosene + charcoal | 184 (46%) |
| Kerosene only | 90 (23%) |
| Charcoal only | 12 (3%) |
| Main lighting fuel |  |
| Electricity/solar | 169 (73%) |
| Kerosene, candles, other | 73 (27%) |
| Number of home meals cooked/day |  |
| 2 | 34 (13%) |
| 3 | 218 (82%) |
| 4+ | 12 (5%) |
| Household size (no. of rooms) |  |
| 1 | 124 (49%) |
| 2 | 68 (27%) |
| 3+ | 59 (24%) |
| No. of household inhabitants |  |
| 1-2 | 58 (14%) |
| 3-4 | 207 (48%) |
| 5-6 | 122 (28%) |
| 7+ | 41 (10%) |
| Female head level of education |  |
| None | 9 (4%) |
| Primary | 101 (40%) |
| Secondary or university | 143 (56%) |
| Male head level of education |  |
| None | 4 (2%) |
| Primary | 57 (26%) |
| Secondary or university | 160 (72%) |
| Sex of main cook |  |
| Female | 211 (83%) |
| Sex of cooking decision maker |  |
| Female | 153 (59%) |

1. 6kg cylinder with on top burner and ring top
2. Portable ceramic charcoal stove commonly used in Kenya

One quarter (27%; n=113) of households reported using a single burner LPG (Meko) stove prior to registering with PayGo Energy, with three-quarters (78%; n=323) using a kerosene stove and over half (57%; n=237) using a charcoal stove (Table 1). Of the 113 customers using LPG prior to registering with PayGo Energy, only half (n=57) exclusively used LPG for cooking. Two PayGo Energy customers that were interviewed mentioned that they continued to use polluting cooking fuels when boiling some foods that require high gas consumption, such as cereals or beans, in order to save money. The overall prevalence of fuel stacking (use of multiple fuels) in the community was 60% (n=239) prior to registering with PayGo Energy.

### 3.2 Pay-as-you-go LPG cooking patterns before COVID-19 lockdown

PayGo Energy residential customers used LPG for cooking an average of 1.4 cooking events/day (SD: 0.5) and four days/week (SD:1.3) (Table 2). Meals prepared using PAYG LPG lasted an average of less than 14 minutes (SD: 13.4). Average PAYG LPG consumption among residential customers over the analysis period (before COVID-19 lockdown) was 0.97 kg/capita/month (11.6 kg/capita/yr) (Table 2), which translated to 3.2 kg of LPG per household per month.

**Table 2.** Pay-As-You-Go (PAYG) LPG fuel usage and spending habits of residential customers before (January 2018-February 2020) and during (March 2020-June 2020) the COVID-19 lockdown **(N=415)**

| Metric | Pre-Lockdown<br>(all months) |  | Pre-Lockdown<br>(January &<br>February only) |  | Pre-Lockdown<br>(June & July only) |  |
| --- | --- | --- | --- | --- | --- | --- |
|  | Mean | SD | Mean | SD | Mean | SD |
| Kg of gas/capita/month | 0.97 | 0.74 | 0.96 | 0.71 | 1.21 | 1.08 |
| Cooking events per day | 1.38 | 0.52 | 1.01 | 0.17 | 1.10 | 0.41 |
| Number of days used/month | 15.6 | 2.0 | 14.0 | 5.6 | 15.9 | 6.8 |
| Number of days used/week | 4.1 | 1.3 | 3.7 | 1.2 | 3.7 | 1.3 |
| Kg of gas used per cooking event<sup>1</sup> | 0.04 | 0.05 | -- | -- | -- | -- |
| Cooking time per event (minutes)<sup>2</sup> | 13.8 | 13.4 | -- | -- | -- | -- |
| Days between payments (median (IQR)) | 8.0 | [4, 22] | 15.0 | [6.0, 30.0] | 10.0 | [5.0, 30.0] |
| Total amount spent per month (KSh) | 840 | 488 | 812 | 361 | 860 | 433 |
| Number of payments/month | 6.1 | 3.3 | 3.1 | 2.1 | 4.3 | 2.9 |
| LPG 6kg cylinders under BCRM: # of payments/month<sup>5</sup> | 0.78 | 0.25 | -- | -- | -- | -- |
| Kerosene: # of payments/month<sup>3</sup> | 17.4 | 13.7 | -- | -- | -- | -- |
| Charcoal: # of payments/month<sup>3</sup> | 22.1 | 12.3 | -- | -- | -- | -- |
| Single payment amount (KSh) | 220 | 212 | 225 | 265 | 184 | 211 |
| Kg of LPG credits purchased per payment | 1.3 | 1.2 | 1.3 | 1.2 | 1.1 | 1.0 |
| LPG 6kg cylinders under BCRM: Single payment amount (KSh)<sup>3</sup> | 988 | 25 | -- | -- | -- | -- |
| Kerosene: Single payment amount (KSh)<sup>3</sup> | 275 | 425 | -- | -- | -- | -- |
| Charcoal: Single payment amount (KSh)<sup>3</sup> | 156 | 315 | -- | -- | -- | -- |

1. Kg of gas used presented from a subset (n=232; 53%) of customers using cylinder smart meters, which records consumption to the nearest 0.001 kg. Usage data from using Goldcard meters (which is rounded to the nearest 0.2 kg) was excluded for accuracy.
2. Data only available from a subset (n=201; 46%) of customers using cylinder smart meters
3. Cross-sectional self-report survey data obtained only at time of customer registration with PayGo Energy (2017)

Average PAYG LPG consumption among commercial customers was much higher (17.1 kg/month) (Table S9); all consumption patterns for the remainder of this analysis focus on residential customers, who make up the vast majority (97%) of PayGo Energy’s clientele.

To assess seasonal differences in LPG usage patterns, PAYG LPG consumption was examined exclusively in two months during the Kenyan hot, dry season (January, February) and the cooler, dry season (June, July) in 2018/2019 (prior to COVID-19 lockdown) (Table 2). Consumption was an average of 0.25 kg/capita/month lower in Kenyan hot, dry season (January-February: 0.96 kg/capita/month) compared with the cooler, dry season (June-July: 1.21 kg/capita/month) (Figure 1). The potential effect of seasonal income fluctuations was documented in a PayGo Energy customer interview:

**Figure 1.**
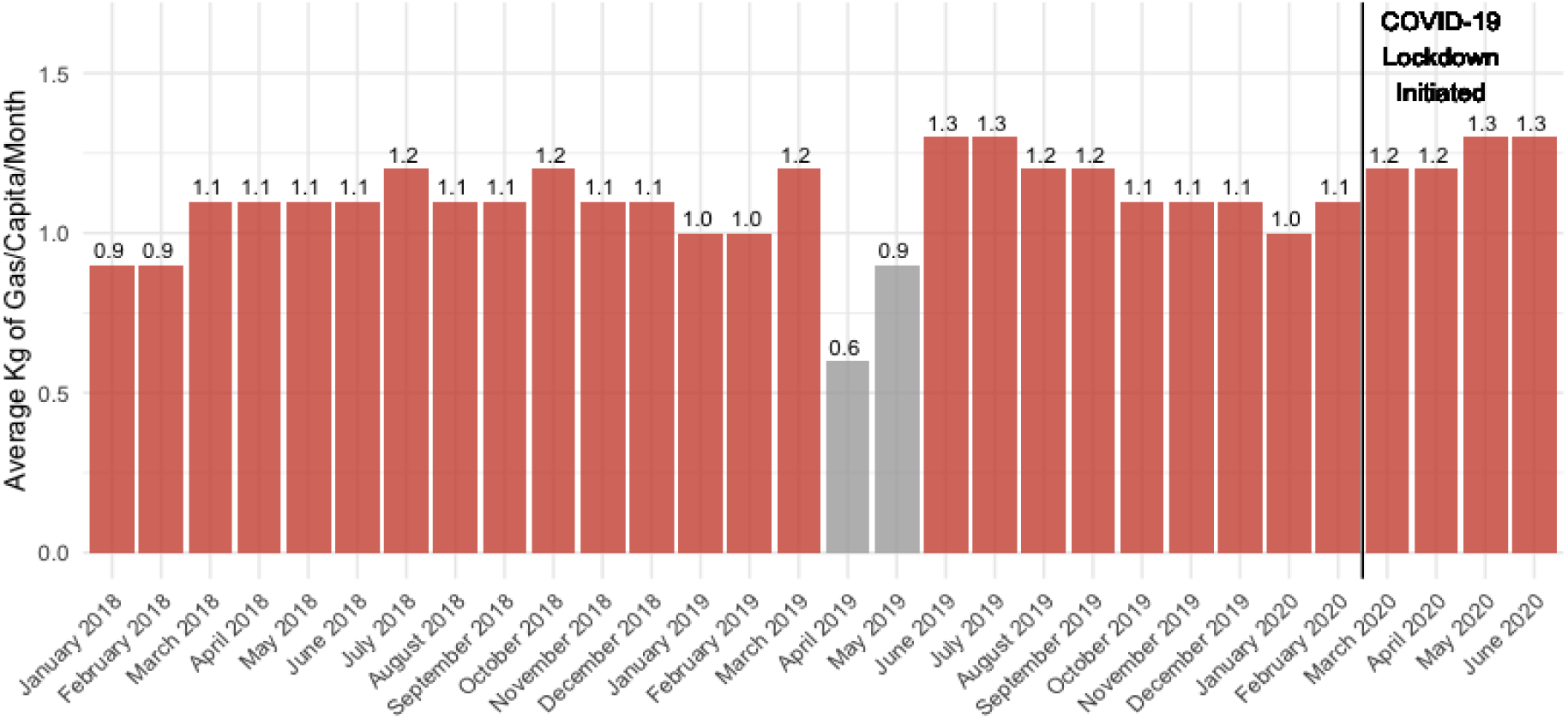
Average kilograms of LPG consumed per month per capita among PayGo Energy residential customers. Note: the greyed bars in April-May 2019 reflect a time period when PayGo Energy updated their LPG metering technology from Goldcard meters to cylinder smart meters and replaced the equipment in several households; data during these months does not reflect true consumption.

> *“Money to top up can be hard to find but you understand that it is just for a season before things revert back to normal.”*

### 3.3 Pay-as-you-go LPG spending patterns

During the analysis period, customers spent an average of 840 Kenyan Shilling (KSh)/ US$7.69 (SD: 488) on PAYG LPG per month, with average single mobile money payments of $220 KSh/ US$2.01 (SD: 212) (Table 2). The mean mobile money payment amounts (220 KSh) were comparable to the average cost of kerosene in the community (275 Ksh) and more than the cost of single charcoal (156 KSh) in 2017. As PayGo Energy charged roughly 180 KSh/ US$1.66 per kilogram of LPG throughout 2018-2020, the mean mobile money payment translated to an average of 1.3 kg (SD: 1.2) of LPG credits purchased (approximately one-quarter of a typical 6 kg gas cylinder).

### 3.4. Pay-as-you-go LPG cooking patterns during COVID-19 lockdown

As the months of PAYG LPG customer data available during COVID-19 lockdown spanned March-June 2020, smart meter data from 2018 and 2019 was restricted to the same months to control for seasonal differences when assessing the impact of COVID-19 lockdown on PAYG LPG cooking behaviors. During lockdown, the average length of a cooking event decreased slightly (from 14.4 minutes to 13.5 minutes).

However, the average number of daily cooking events during lockdown (1.72) was 60% higher than that of the same months in 2018/2019 (1.07) (Figure 2). Further, the mean number of days per week using PAYG LPG increased from 4.3 (pre-lockdown) to 5.0 (lockdown) (p<0.001). This resulted in an overall increase in PAYG LPG consumption during lockdown, from 1.12 kg/capita/month (SD: 0.75) to 1.25 kg/capita/month (SD: 1.01). Five out of six residential customers interviewed affirmed that their use of PAYG LPG during the lockdown increased due to having to prepare lunch while their children were home from school; children ordinarily eat their lunch at school.

**Figure 2.**
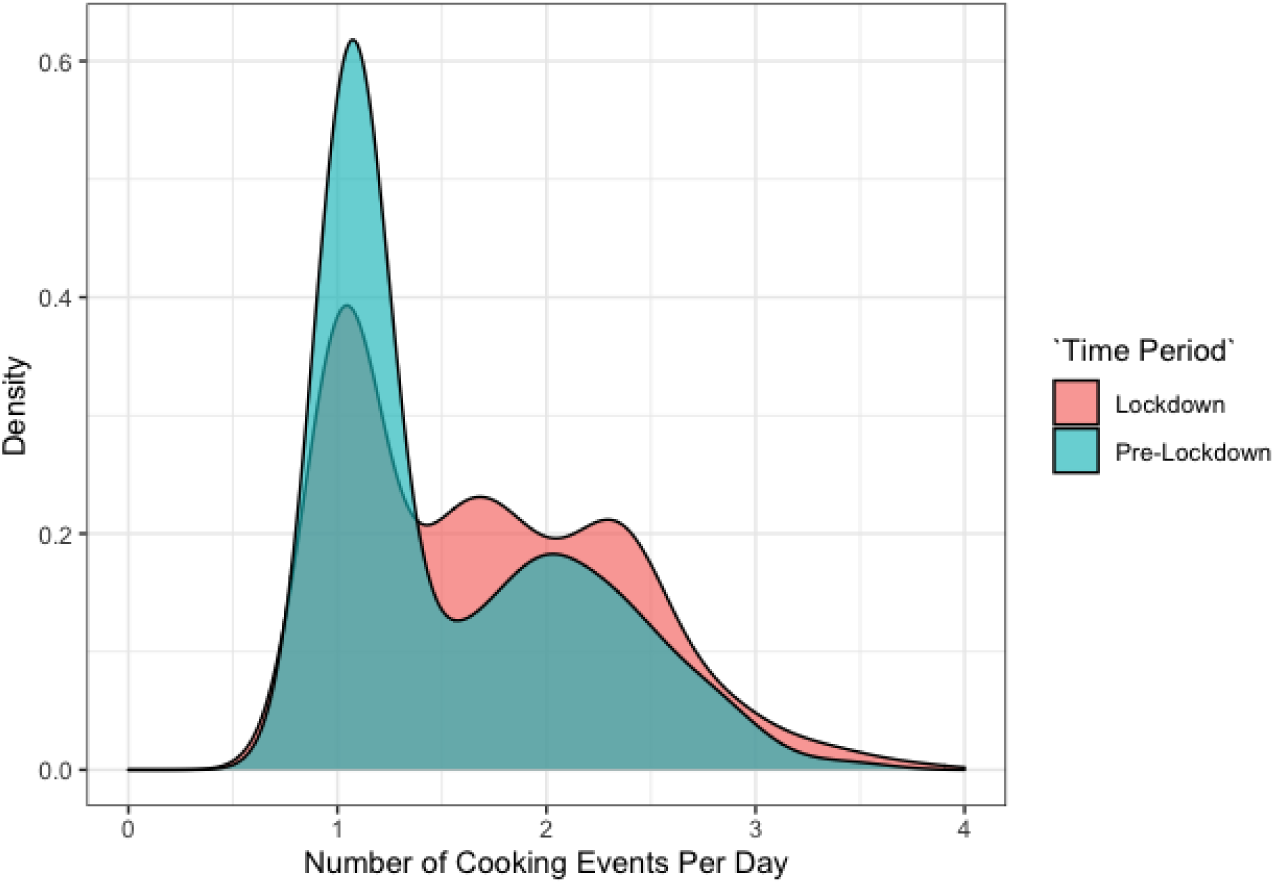
Number of cooking events per day using PAYG LPG stove before (‘Pre-Lockdown) and during COVID-19 community lockdown (‘Lockdown’)

### 3.5. Pay-as-you-go LPG spending patterns during COVID-19 lockdown

Among households using PAYG LPG during the lockdown, the average single payment amount significantly dropped by nearly 50% (from 336 to 179 KSh/ US$3.08 to US$1.64) (p<0.001) compared with payments made during March-June of 2018/2019 (Figure 3a). This resulted in the mean amount of LPG purchased per payment shifting from 2 kg (pre-lockdown) to just over 1 kg (lockdown) (Table 3). Lower payments during lockdown (Figure 3a) were offset by a 67% increase in median payment frequency - from 4.6 payments/month (SD: 3.2) to 7.7 (SD: 1.9) payments/month during lockdown (Table 3). This resulted in customers making mobile money payments every 4 days (IQR:[2.5, 9.0]) during COVID-19 lockdown, compared with once every 8 days (IQR:[4.0, 22.0]) pre-lockdown (Figure 3b). This change in payment patterns results in an insignificant decrease (p=0.29) in total PAYG LPG monthly expenditure (867 Ksh/ $7.94 USD (pre-lockdown) to 816 Ksh/ $7.47 USD (lockdown)).

**Table 3.**
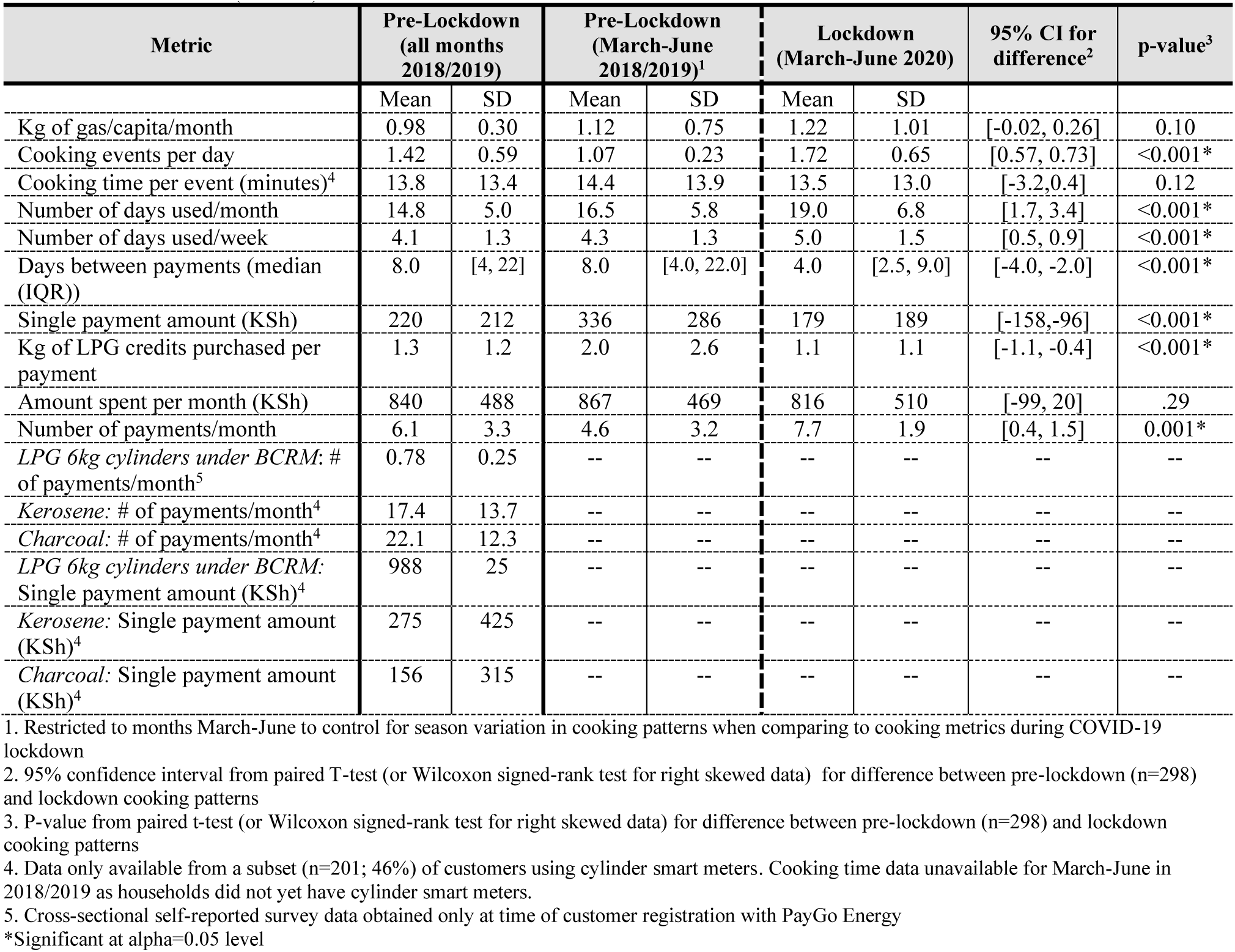
Pay-As-You-Go (PAYG) LPG fuel usage and spending habits of residential customers active during the COVID-19 lockdown (N=298)

| Metric | Pre-Lockdown<br>(all months<br>2018/2019) |  | Pre-Lockdown<br>(March-June<br>2018/2019)<sup>1</sup> |  | Lockdown<br>(March-June 2020) |  | 95% CI for<br>difference<sup>2</sup> | p-value<sup>3</sup> |
| --- | --- | --- | --- | --- | --- | --- | --- | --- |
|  | Mean | SD | Mean | SD | Mean | SD |  |  |
| Kg of gas/capita/month | 0.98 | 0.30 | 1.12 | 0.75 | 1.22 | 1.01 | [-0.02, 0.26] | 0.10 |
| Cooking events per day | 1.42 | 0.59 | 1.07 | 0.23 | 1.72 | 0.65 | [0.57, 0.73] | <0.001* |
| Cooking time per event (minutes)<sup>4</sup> | 13.8 | 13.4 | 14.4 | 13.9 | 13.5 | 13.0 | [-3.2, 0.4] | 0.12 |
| Number of days used/month | 14.8 | 5.0 | 16.5 | 5.8 | 19.0 | 6.8 | [1.7, 3.4] | <0.001* |
| Number of days used/week | 4.1 | 1.3 | 4.3 | 1.3 | 5.0 | 1.5 | [0.5, 0.9] | <0.001* |
| Days between payments (median<br>(IQR)) | 8.0 | [4, 22] | 8.0 | [4.0, 22.0] | 4.0 | [2.5, 9.0] | [-4.0, -2.0] | <0.001* |
| Single payment amount (KSh) | 220 | 212 | 336 | 286 | 179 | 189 | [-158, -96] | <0.001* |
| Kg of LPG credits purchased per<br>payment | 1.3 | 1.2 | 2.0 | 2.6 | 1.1 | 1.1 | [-1.1, -0.4] | <0.001* |
| Amount spent per month (KSh) | 840 | 488 | 867 | 469 | 816 | 510 | [-99, 20] | .29 |
| Number of payments/month | 6.1 | 3.3 | 4.6 | 3.2 | 7.7 | 1.9 | [0.4, 1.5] | 0.001* |
| LPG 6kg cylinders under BCRM: #<br>of payments/month<sup>5</sup> | 0.78 | 0.25 | -- | -- | -- | -- | -- | -- |
| Kerosene: # of payments/month<sup>4</sup> | 17.4 | 13.7 | -- | -- | -- | -- | -- | -- |
| Charcoal: # of payments/month<sup>4</sup> | 22.1 | 12.3 | -- | -- | -- | -- | -- | -- |
| LPG 6kg cylinders under BCRM:<br>Single payment amount (KSh)<sup>4</sup> | 988 | 25 | -- | -- | -- | -- | -- | -- |
| Kerosene: Single payment amount<br>(KSh)<sup>4</sup> | 275 | 425 | -- | -- | -- | -- | -- | -- |
| Charcoal: Single payment amount<br>(KSh)<sup>4</sup> | 156 | 315 | -- | -- | -- | -- | -- | -- |

1. Restricted to months March-June to control for season variation in cooking patterns when comparing to cooking metrics during COVID-19 lockdown
2. 95% confidence interval from paired T-test (or Wilcoxon signed-rank test for right skewed data) for difference between pre-lockdown (n=298) and lockdown cooking patterns
3. P-value from paired t-test (or Wilcoxon signed-rank test for right skewed data) for difference between pre-lockdown (n=298) and lockdown cooking patterns
4. Data only available from a subset (n=201; 46%) of customers using cylinder smart meters. Cooking time data unavailable for March-June in 2018/2019 as households did not yet have cylinder smart meters.
5. Cross-sectional self-reported survey data obtained only at time of customer registration with PayGo Energy
\*Significant at alpha=0.05 level

**Figure 3.**
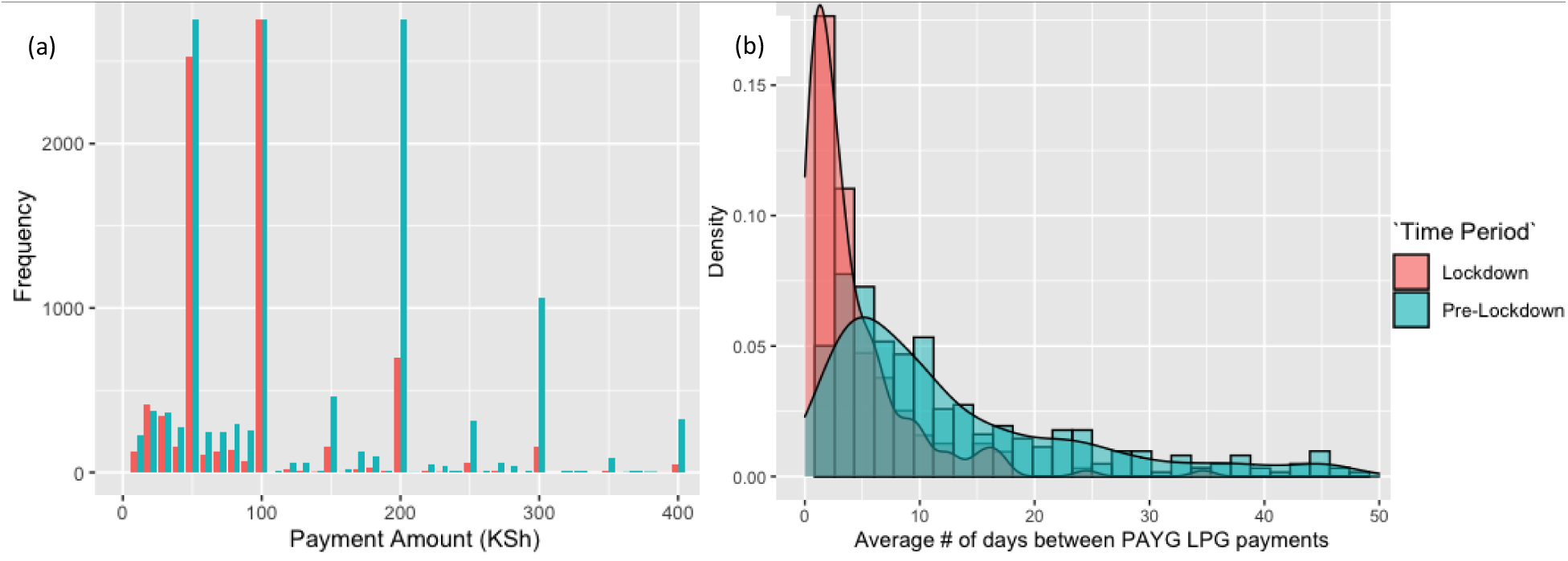
(a) Single mobile money payments made for PAYG LPG before (‘Pre-Lockdown’) and during COVID-19 community lockdown (‘Lockdown’) (b) Number of days between subsequent PAYG LPG payments before (‘Pre-Lockdown) and during COVID-19 community lockdown (‘Lockdown’)

### 3.6 Pay-as-you-go LPG consumption by socioeconomic characteristics

Per capita consumption among customers that purchased full LPG refills under the BCRM before registering for PAYG LPG substantially increased by 0.35 kg/month during the lockdown (from 0.88 (SD: 0.17) to 1.23 (SD: 0.05)) (Table 4). PAYG LPG monthly per capita consumption increased by less than half that of previous LPG users (0.15 kg/capita/month) among households that had not used LPG before PAYG LPG registration during the lockdown (from 1.12 (SD: 0.24) to 1.27 (SD: 0.03). PAYG LPG per capita consumption during the lockdown increased by approximately 0.4 kg/capita/month during the lockdown among households where the head female was employed (in the formal or informal sector), while only slightly increasing (0.1 kg/capita/month) among those employed in casual jobs and decreasing substantially among those unemployed before lockdown (−0.88 kg/capita/month) (Table 4). Average PAYG LPG monthly consumption also increased among households with three or more family members, while decreasing among those with one or two family members. The reason for this dichotomy is likely having to prepare lunch children home from school during lockdown.

**Table 4.** Monthly Per Capita PAYG LPG consumption by pre-registration fuel choice and occupation (N=277)

|  | N | Monthly Kg Per Capita Usage (Mean (SD)) |  | Cooking Time Per Day (Minutes)* (Mean (SD)) |  | Single payment amount (KSh) (Mean (SD)) |  | Total Spent per month (KSh) (Mean (SD)) |  |
| --- | --- | --- | --- | --- | --- | --- | --- | --- | --- |
|  |  | Pre-Lockdown (March - June 2018 & 2019) | Lockdown (March - June 2020) | Pre-Lockdown (All months 2018 & 2019) | Lockdown (March - June 2020) | Pre-Lockdown (March - June 2018 & 2019) | Lockdown (March - June 2020) | Pre-Lockdown (March - June 2018 & 2019) | Lockdown (March - June 2020) |
| LPG user prior to PAYG LPG |  |  |  |  |  |  |  |  |  |
| Yes | 91 | 0.88 (0.17) | 1.23 (0.05) | 62 (26) | 65 (30) | 304 (285) | 180 (232) | 820 (77) | 793 (42) |
| No | 186 | 1.12 (0.24) | 1.27 (0.03) | 80 (72) | 81 (65) | 346 (297) | 175 (180) | 870 (50) | 832 (33) |
| Primary fuels prior to PAYG LPG |  |  |  |  |  |  |  |  |  |
| Kerosene only | 64 | 1.14 (0.26) | 1.32 (0.06) | 64 (32) | 67 (34) | 331 (307) | 159 (164) | 789 (63) | 858 (46) |
| Charcoal only | 11 | 1.25 (0.90) | 1.34 (0.10) | 194 (154) | 158 (157) | 238 (150) | 132 (89) | 404 (162) | 576 (123) |
| Kero. + charcoal | 111 | 1.11 (0.24) | 1.22 (0.05) | 68 (39) | 70 (34) | 351 (299) | 187 (195) | 892 (59) | 828 (25) |
| LPG only | 50 | 1.30 (0.34) | 1.33 (0.07) | 61 (22) | 66 (26) | 351 (388) | 196 (282) | 770 (174) | 770 (48) |
| LPG + Kerosene | 18 | 0.74 (0.20) | 1.12 (0.07) | 65 (37) | 52 (35) | 180 (105) | 146 (182) | 707 (178) | 752 (43) |
| LPG + Charcoal | 8 | 1.03 (0.35) | 1.39 (0.17) | 81 (15) | 73 (42) | 218 (122) | 165 (156) | 1077 (216) | 1160 (46) |
| LPG + Charcoal + Kerosene | 15 | 0.76 (0.15) | 0.98 (0.08) | 45 (19) | 46 (25) | 354 (309) | 183 (123) | 857 (121) | 742 (82) |
| Occupation (female) |  |  |  |  |  |  |  |  |  |
| Formal job | 7 | 1.12 (0.26) | 1.52 (0.11) | 140 (11) | 132 (25) | 207 (109) | 141 (66) | 1068 (90) | 1142 (49) |
| Informal sector job | 23 | 1.03 (0.29) | 1.39 (0.08) | 70 (46) | 61 (35) | 390 (343) | 248 (274) | 832 (61) | 941 (49) |
| Day labor/casual job | 13 | 0.99 (0.24) | 1.09 (0.1) | 102 (40) | 83 (25) | 263 (357) | 166 (118) | 779 (193) | 815 (108) |
| Unemployed | 2 | 1.50 (0.63) | 0.62 (0.08) | -- | -- | 560 -- | 100 -- | 1516 -- | 1110 -- |
| No. people in home |  |  |  |  |  |  |  |  |  |
| 1-2 | 58 | 2.22 (0.47) | 2.14 (0.25) | 47 (24) | 49 (38) | 331 (307) | 159 (195) | 843 (76) | 706 (63) |
| 3-4 | 201 | 1.09 (0.24) | 1.29 (0.02) | 69 (44) | 71 (38) | 347 (315) | 183 (230) | 821 (56) | 783 (40) |
| 5-6 | 118 | 0.78 (0.16) | 0.91 (0.03) | 81 (59) | 82 (64) | 310 (241) | 168 (136) | 840 (41) | 854 (56) |
| 7+ | 41 | 0.56 (0.13) | 0.68 (0.06) | 119 (114) | 113 (94) | 361 (332) | 176 (138) | 981 (80) | 1079 (36) |

\*Cooking time per day only available for a subset (n=201; 46%) of PayGo Energy customers with cylinder smart meters. Data provided for all months in 2018/2019 as cylinder smart meters weren't installed until after June 2019.
Note: some demographic data only collected from a subset of customers during certain years of enrollment by PayGo Energy (sample size varies by variable). Numbers for certain variables do not sum up to overall sample size due to missing data. Male occupation not presented due to low sample size.

Similarly, monthly expenditure on PAYG LPG was higher during COVID-19 lockdown among households with the female household head employed in formal or informal sectors compared with a decrease in expenditure in households where the female head was unemployed or worked as a day laborer. Total monthly amount spent on PAYG LPG increased among households with five or more family members, while decreasing among families with four members or less (Table 4).Single payment amounts decreased by ∼50% among households not previously purchasing full cylinder LPG while decreasing only 40% among households using LPG prior to registering with PayGo Energy (Table 4). Notably, average monthly expenditure on PAYG LPG during lockdown increased by the largest margin among households previously using charcoal before registering with PayGo Energy. One interviewed customer attributed their higher use of PAYG LPG to higher prices of polluting fuels in the community:

> *“Gas is more economical compared to kerosene or charcoal […] gas prices are affordable and [PAYG LPG] is sustaining us now.”*

### 3.7 Changes in pay-as-you-go LPG consumption with user experience

PAYG LPG monthly per capita consumption gradually increased overtime among PayGo Energy customers (Table 5). In the first 6 months since registering with PayGo Energy, average customer consumption was 0.76 kg/capita/month; monthly per capita consumption increased to

**Table 5.** Long-term consumption patterns by length of time as a PAYG LPG customer (among households that registered with PayGo Energy in 2017 (N=207*).

| Number of months since PAYG LPG account activation | N | Kg of gas/ capita/month |  | Number of days used/month |  | Cooking events per day |  | Amount spent per month (KSh) |  |
| --- | --- | --- | --- | --- | --- | --- | --- | --- | --- |
|  |  | Mean | SD | Mean | SD | Mean | SD | Mean | SD |
| 1-6<sup>1</sup> | 125 | 0.76 | 0.63 | 12.4 | 5.3 | 1.03 | 0.11 | 735 | 437 |
| 7-12 | 181 | 0.96 | 0.67 | 14.3 | 5.6 | 1.05 | 0.04 | 791 | 458 |
| 13-18 | 207 | 0.98 | 0.72 | 13.9 | 5.7 | 1.05 | 0.08 | 802 | 526 |
| 19-24 | 196 | 1.09 | 0.99 | 12.9 | 5.6 | 1.07 | 0.08 | 820 | 553 |

\*This sample size includes only customers recruited in ‘Wave 1’ (i.e. in 2017) to examine cooking patterns unimpacted by COVID-19 lockdown. This example excludes households that deactivated their account within two years of registration to examine long-term cooking behaviors. Smart meter data from January and February were excluded from this analysis to control for seasonal cooking pattern differences confounding the relationship.
1. The sample size for months 1-6 is smaller as 82 households recruited in Wave 1 prior to September 2017 were not initially installed with a smart meter, so no data is available. A sensitivity analysis with these 82 households excluded revealed no significant changes in long-term consumption patterns.

0.96 kg/capita month after households had been PAYG LPG customers for at least 6 months. The increase in monthly consumption was complemented by an increase in monthly expenditure by nearly 100 KSh between the first and second year as a PAYG LPG customer, from an average of 735 Ksh ($US6.75) spent per month in months 1-6 to 820 KSh (US$7.53) in months 19-24.

Customers that did not deactivate their PayGo Energy account in 2018/2019 had a higher proportion (61%) of female household members in charge of cooking fuel decisions than males, compared with households that deactivated their PAYG LPG account during this period (48%) (Table 6). Additionally, the female household head was significantly more likely (p=0.04) to be employed in a formal sector job and less likely to be unemployed among households keeping their PAYG LPG account active, compared with customers that deactivated their PAYG LPG account in 2018/2019. There was no association between (Meko) LPG stove use prior to registering with PAYG LPG and customers maintaining or deactivating their PayGo Energy account during the study period.

**Table 6.**
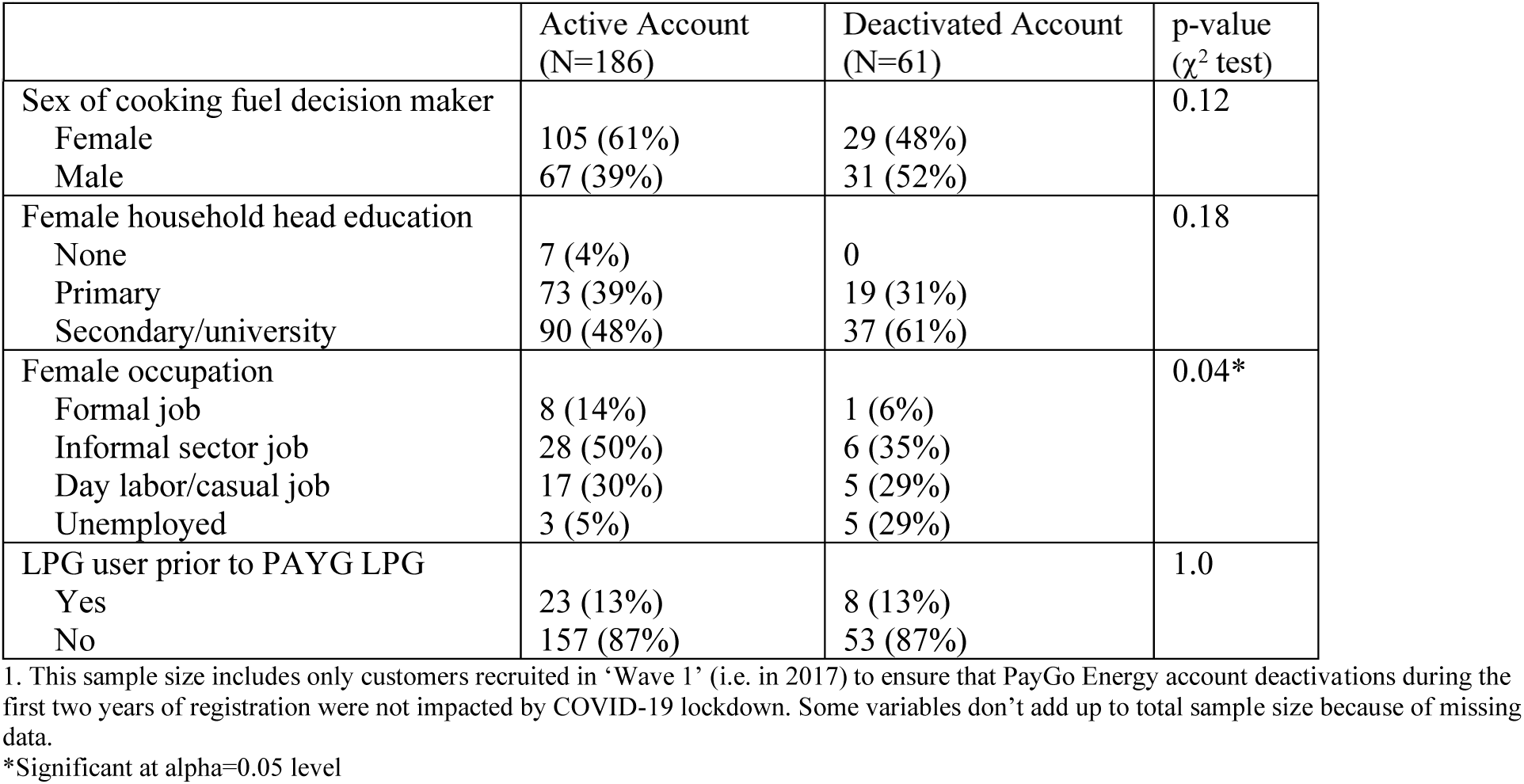
Factors associated with PAYG LPG account deactivations in 2018/2019 (before COVID-19 lockdown) (N=247^1^)

### 3.8 Perceived benefits of pay-as-you-go LPG versus traditional full cylinder LPG and kerosene use

PAYG LPG was revealed to have a number of benefits compared with purchasing a full 6kg cylinder of LPG and/or kerosene. Benefits reported by seven interviewed participants included fuel affordability, safety from burns/gas explosions, time savings, ease of fuel access and user-friendliness of the smart meters. The ability to make small, regular payments was critical to participants being able to cook with LPG, who were reassured to know that they could still prepare meals for their children with only small amounts of money at their disposal:

> *“Because even if you have the lowest amount of money, you will still be able to cook.” “It is okay because if I have even 20 KSh, I can refill [make a mobile money payment] and finish cooking.”*

For some customers who already had access to full cylinder LPG before registering with PayGo Energy, PAYG LPG often serves as a back up fuel source when they may have less cash on hand:

> *“[PAYG LPG] has helped me in such a way that if I don’t have enough money to fill the bigger cylinder, I can get PayGo.”*

The value for money and efficiency in terms of the amount of meals that could be prepared was another commonly reported benefit discussed by PayGo Energy customers when interviewed:

> “*PayGo is very economical because when I was buying kerosene, I would pay 100 KSh and it would not be enough for my cooking. I could not boil water, use for bathing and cook supper with [a single purchase of] kerosene*…*I would not have any [fuel] left to cook breakfast in the morning. Now when I refill gas [using PAYG LPG] for 100 KSh, I use it for all those things including breakfast the next day without straining.”*

Multiple first time LPG users also favored the technology itself. They enjoyed the efficiency of cooking of the double-burner LPG stove, which they reported saves time by enabling them to cook dishes (e.g. ugali and vegetables) simultaneously on each burner. PAYG LPG was also considered to be more convenient because participants did not have to carry the cylinder to the station for refilling, nor did they have to buy matchsticks to start the flame. Another interviewee pointed out the benefits of using the mobile money system (M-Pesa) for purchasing LPG credits, as other community members paying for cooking fuels via a ‘card system’ are sometimes unable to cook if the card is misplaced.

Increased safety was also a reported advantage of PAYG LPG as several customers interviewed were worried about gas explosions and risks of their children being burned if purchasing full cylinder LPG. Many interviewees stated they had greater peace of mind cooking with PAYG LPG because “when there is a leakage [PayGo Energy customer support] can detect from the main office and shut off [the gas].” One interviewee indicated that they previously refrained from buying full cylinder 6kg LPG (and used kerosene instead) because a friend in the community suffered a burn when cooking with gas; had it not been for the safety of smart meter technology of PAYG LPG, this customer would not be cooking with LPG until their children were older and could be trusted to keep their distance from the cookstove and cylinder.

LPG was viewed as an aspirational fuel for the interviewees: “*I never had the hope of one day owning an LPG [stove] in my home*”. Another customer stated: “*And by the way, I have seven children and I [am able to use] LPG*” because of PAYG LPG. Indeed, increased monthly consumption and expenditure of PAYG LPG among households with seven or more members during lockdown (Table 4) suggests that LPG was able to accommodate higher cooking needs due to several children being home from school.

### 3.9 Stove use monitoring data in Eldoret, Kenya

Study households in Eldoret, Kenya had statistically significantly higher SES than PayGo Energy customers in Mukuru kwa Reuben by all factors examined (education, occupation, household size, and electricity access for lighting) (Table S4). Despite higher SES in Eldoret, PAYG LPG companies have not yet penetrated peri-urban communities in Kenya due to higher infrastructure costs of distributing outside large cities. Thus, LPG in Eldoret is currently only available through full filled cylinder purchase (6 kg cylinders are the most common size).

Among seven households from Eldoret with available SUM data and primarily cooking with LPG, the number of days/month using the cooking fuel declined by 75% during the COVID-19 lockdown, from an average of 17 days in March 2020 to four days or less in April-May 2020 (Figure 4). In contrast, households primarily cooking with charcoal/wood (n=16) continued to use these fuels consistently during lockdown, approximately 20 days per month (Figure 4).

**Figure 4.**
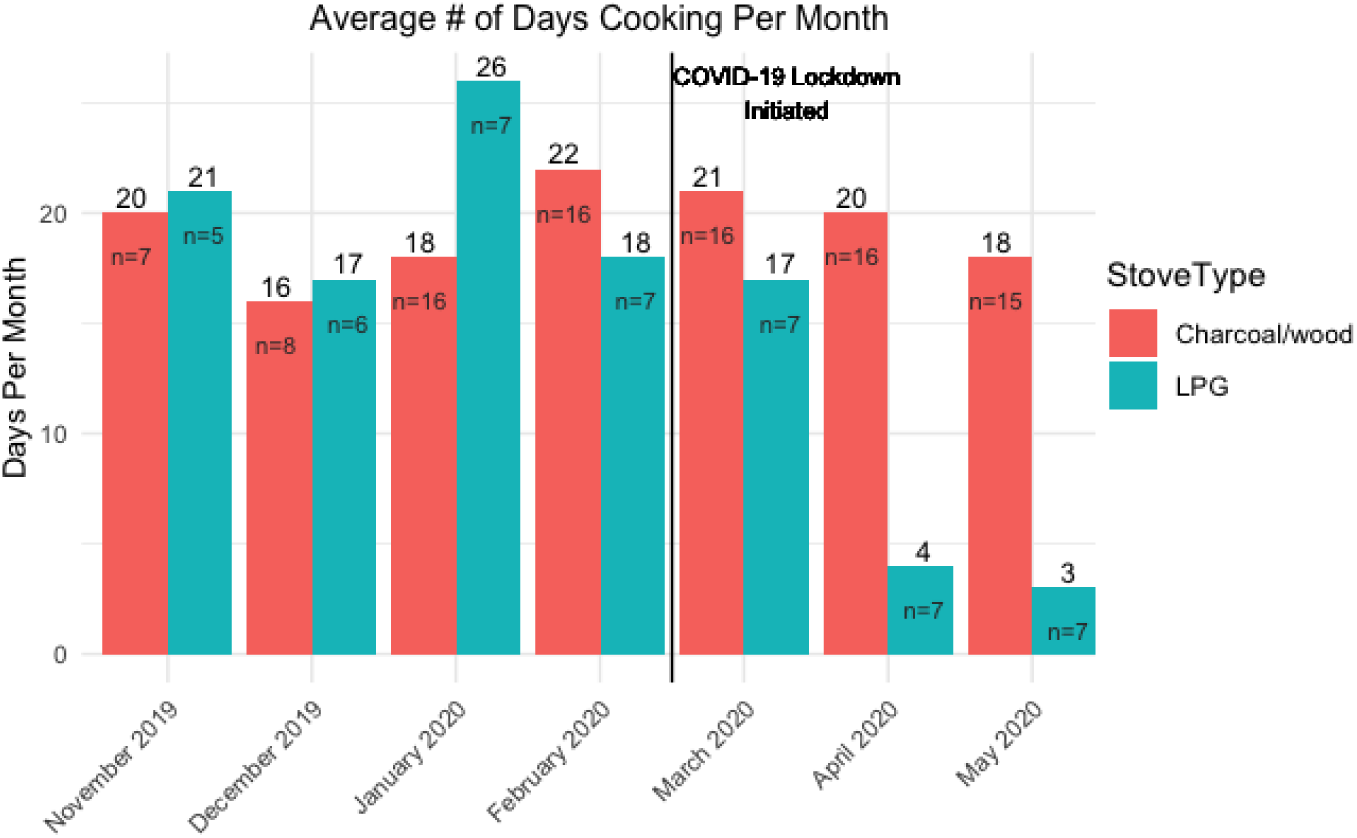
Average number of days per month cooking with LPG versus traditional stoves in Eldoret, Western Kenya before and during a COVID-19 lockdown measured with stove use monitors.

The average duration of cooking per day (hours) using charcoal and wood stoves increased by 200-300% in Eldoret during lockdown (e.g. wood stove daily cooking time increased from 2 to 5.5 hours/day) (Figure 5a). Conversely, among those who continued to use LPG (n=5), daily cooking time decreased slightly during lockdown (Figure 5b).

**Figure 5.**
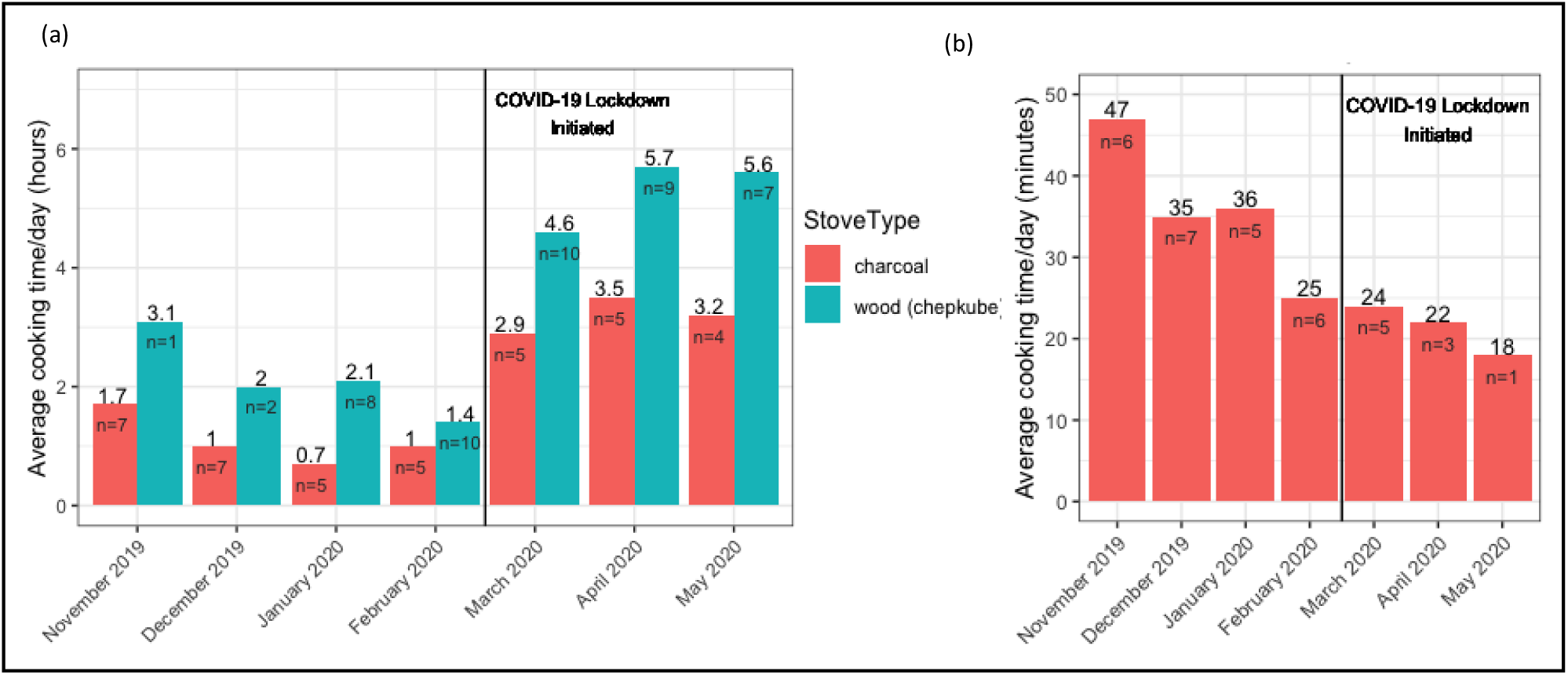
Average hours per day per month cooking on (a) wood and charcoal stoves and (b) LPG stoves in Eldoret, Kenya before and during a COVID-19 lockdown.

## 4. Discussion

This study is the first to report long-term cooking and spending patterns in East Africa using PAYG LPG smart meter data. This study also uniquely evaluated use of PAYG LPG in the context of economic instability introduced by COVID-19 control measures. Among 415 PAYG LPG residential customers, annual LPG per capita consumption was 11.6 kg/capita/year, below that of peri-urban dwellers (12.8 kg/capita/year) and of urban dwellers (18.7 kg/capita/year) reported in the 2019 Kenya LPG National Feasibility study of domestic fuel patterns.^41^ The mean payment made by PayGo Energy customers (220 KSh) was similar to the typical amount spent for kerosene in the community (275 KSh), and approximately one-fourth of what one would typically pay for a full 6kg cylinder of LPG (988 KSh), suggesting that households prefer to pay for gas in smaller, more frequent increments than required for the full LPG cylinder. On a monthly basis, the average amount spent by PAYG LPG residential customers (840 Ksh/month (US$7.69)) was similar to the paid amount reported by purchasers of LPG cylinder refills under the BCRM in the same community (850 Ksh/month (US$7.83)).^32^ Similar average monthly expenditures among PAYG LPG customers despite lower usage of PAYG LPG (4 days/week) (Table 2) compared to self-reported use of LPG under the BCRM (6.3 days/week)^32^ is due to a ∼7.1% fuel surcharge by PayGo Energy to cover equipment installation and delivery fees.^42,43^

Additional to the flexible payment scheme offered by PAYG LPG, in interviews with customers it was revealed that increased safety from burns/gas explosions, time savings and ability to prepare separate dishes on both burners and fuel delivery were also key features enjoyed by households. The participants detected that they were able to carry out a higher amount of cooking/bathing tasks with PAYG LPG compared with the same amount spent on kerosene.

While four of the six residential households stated in interviews that they used PAYG LPG to meet all of their cooking needs (with the exception of electricity (e.g. electric jug) to warm water for bathing), two study customers that were interviewed indicated that they continued to cook with kerosene or charcoal alongside PAYG LPG. They used polluting fuels for boiling foods such as cereals or beans to save money, as they believe they would not be able to afford the amount of gas required to prepare food items that required a prolonged cooking time.

Cooking fuel stacking (concurrent use of multiple clean and polluting fuels) by the study population may be a primary cause of lower annual per capita consumption among PAYG LPG users relative to those purchasing the full LPG cylinder. Fuel stacking was high (60%) among LPG users buying full cylinder refills prior to using PAYG LPG (Table 1). As PAYG LPG consumption during the lockdown notably decreased among those employed in casual jobs or unemployed (Table 4), the inability to afford exclusive use of LPG under a PAYG scheme may lead to stacking with polluting fuels for some resource-poor households. Nonetheless, five of the seven PayGo Energy customers interviewed indicated that they would be cooking kerosene had they not was not had access to PAYG LPG. Thus, the ability to pay in smaller payments did provide families that aspired to cook with LPG access to the fuel.

### 4.1 Impacts of COVID-19 lockdown on pay-as-you-go LPG cooking behaviors

Despite financial hardships imposed by the COVID-19 lockdown (95% of households reported decreases in household income during lockdown^32^), an increase in monthly per capita PAYG LPG consumption was observed among residential customers (from 1.12 kg/capita/month pre-lockdown to 1.22 kg/capita/month during lockdown) (Table 3), and 95% (n=288) of active PayGo Energy customers continued to use PAYG LPG during the COVID-19 lockdown (March-June 2020). A rise in LPG consumption during lockdown was likely the result of children staying at home during the day due to schools being closed (children consumed lunch at home when they would ordinarily eat this meal at school); children being home from school was reported by five out of six interviewed PayGo Energy residential customers as the reason for their increase in PAYG LPG usage.

While only 5% of PayGo Energy customers discontinued their use of PAYG LPG during the COVID-19 lockdown, a previous longitudinal survey conducted in the same community revealed that 27% of households using LPG under the BCRM as a primary cooking fuel reverted to purchasing kerosene (14%) or gathering wood for free (13%) during lockdown.^32^ Further, the same study found that four (67%) of six households using full cylinder LPG with a greater number of people to cook for during lockdown switched to wood for cooking. This proportion is much greater than the 22% (10 out of 44) of households with no change in the number of household members during lockdown that switched from full cylinder LPG to wood or kerosene for cooking.^32^ Moreover, one-third (n=2 of 6) of households in Mukuru kwa Reuben cooking with kerosene that had to cook for more household inhabitants during lockdown switched to wood (which they could obtain for free on the side of the road), compared with 8% (n=8 of 97) of kerosene users that reported no changes in family size switching to wood or charcoal for cooking. Thus, increases in LPG consumption among PayGo Energy customers that occurred by having to cook more frequently for more individuals (e.g. children that stayed home from school) may not have occurred had they been purchasing LPG under the BCRM.

The juxtaposition of LPG cooking habits between PayGo Energy customers and full cylinder LPG users within the same community supports the utility of the flexible payment schedule (e.g. smaller amounts more frequently) provided by PAYG LPG, which potentially allowed customers to maintain use of LPG despite income declines during COVID-19 lockdown (88% of community members reported complete cessation of income during lockdown^32^). Access to PAYG LPG may have therefore averted a possible rise in community-level household air pollution emissions from increased use of wood/kerosene for cooking. This is particularly significant considering that three-quarters of PayGo Energy customers were first time LPG users (Table 1).

It is unlikely that higher SES among PayGo Energy customers accounted for greater LPG usage during the lockdown compared with full cylinder LPG users; while a sensitivity analysis revealed that PayGo Energy customers had slightly larger household size and higher prevalence of LPG usage under the BCRM (48%) (at the time of registration for PayGo Energy), compared with the community-level average (37%), female occupation and household education levels were similar between both groups (Table S3). Moreover, individuals living in Eldoret that were more affluent than PayGo Energy customers in Mukuru kwa Reuben (Table S4) reduced their use of full cylinder refill LPG (days/month) by 75%, providing further evidence of the value of PAYG LPG (and not higher SES) in preventing a reversion to polluting cooking fuels during lockdown.

Modifications to cooking patterns among PAYG LPG users were complemented by alterations in the rate of mobile money payments. The median number of days between subsequent payments was reduced by half (from once every eight days pre-lockdown to once every four days during lockdown) (Figure 3b). However, the average payment amount decreased by nearly half (from 336 Kenyan Shillings (KSh)/US$3.08 (pre-lockdown) to 179 KSh/US$1.64 (lockdown)) (Table 3). Despite making a higher volume of payments in smaller increments during the lockdown because of declines in household income due to COVID-19 control measures, customers’ total monthly PAYG LPG expenditure remained relatively constant (867 Ksh/ $7.94 USD (pre-lockdown) to 816 Ksh/ $7.47 USD (lockdown)).

### 4.2 Socioeconomic factors associated with changing cooking and payment behaviors during COVID-19 lockdown

The increase in PAYG LPG per capita consumption and monthly expenditure during lockdown was driven by households with a higher number of family members (children staying home from school); households with at least three family members increased their PAYG LPG consumption during lockdown, while households with two people or less reduced their LPG consumption.

Customers that previously used LPG prior to registering with PayGo Energy increased their PAYG LPG monthly consumption by twice as much (0.35 kg/capita/month) as customers that had not previously used full cylinder LPG (0.15 kg/capita/month) (Table 4). Higher SES among previous LPG users may also partially explain these observed differences; female head of households with jobs in the formal or informal sector increased their PAYG LPG monthly spending during lockdown by 80-90 KSh/month, while spending among those with casual jobs increased by 35 KSh and decreased among those that were unemployed. Monthly spending on PAYG LPG increased most dramatically among households only cooking with charcoal or LPG purchased through a full cylinder in addition to charcoal prior to the lockdown (Table 4); this may be due to a national ban on the sale of charcoal and its movement out of Kenya that was enacted in 2019 to reduce deforestation,^40^ having driven up the price of charcoal in the community.

### 4.3 Seasonality

When restricting smart meter data from 2018-2019 (pre-lockdown) to four specific months that fell in hot or cool season, monthly consumption and expenditure on PAYG LPG was lower in January and February relative to June and July (Table 2). Household incomes are typically lower during the start of the year in this community due to higher expenditure over the December holiday period and school-related expenses that come due in January. These seasonal LPG usage fluctuations, however, are less extreme than what has been observed in India, with LPG cylinder refill sales 10% lower in ‘non-cropping’ season than during cropping/harvest seasons when families typically have higher income.^24^ As PAYG LPG allows consumers to adjust their payment amount based on their financial situation, it may be useful to expand this model to other countries where income changes seasonally.

### 4.4 Customer experience and retention

Average PAYG LPG monthly per capita consumption also gradually increased after registering with PayGo Energy, from 0.76 kg/capita/month (SD: 0.63) in months 1-6 to 1.09 kg/capita/month in months 19-24 (Table 5). The reason for the increase is likely partially a result of lower consumers of PAYG LPG deactivating their account in 2018 or 2019. Households that did not deactivate their PayGo Energy account during the study period had a higher percentage of female cooking fuel decision-makers and female household heads employed in the formal or

informal sector compared with households that deactivated their account (Table 6). This information may be useful for future commercial companies seeking to target the ‘early adopters’ of PAYG LPG technology and ensure continuity of their customer base.

### 4.5 Female Empowerment

The important role of gender equity in energy poverty is well-documented in the literature.^44–46^ Prior to registering for PAYG LPG, females decided use of household cooking fuels at a 20% higher rate in households already using LPG 6kg cylinders (77%), compared with households not using LPG (55%). Among households registering to use PAYG LPG, the proportion of females making decisions about cooking fuel purchases (59%) was significantly higher than the proportion in the community that did not register for PAYG LPG (47%) (Table S3). Further, female participants working in the informal sector or those with ‘casual’ jobs used their PAYG LPG stove significantly less than female participants in formal employment both before and during the lockdown (Table 3). Lastly, the proportion of households with female cooking fuel decision-makers was higher households that did not deactivate their PayGo Energy account prior to the pandemic (61%) compared with households that deactivated their account (48%).

Thus, policies that foster fiscal and social empowerment of women may have important co-benefits in fostering the transition to clean cooking.^47^ These policies are especially important during COVID-19, as female informal sector workers are likely to be among the first to lose their job and livelihoods during a lockdown,^48^ potentially resulting in additional health burden (e.g. poor mental health).^49^

### 4.6 Strengths and Limitations

With a sample of 415 residential PayGo Energy customers, this study was powered to examine cooking behaviours before and during a nationwide lockdown in Kenya. As customers were recruited primarily through door-to-door advertising, PayGo Energy staff members identifying a new customer (e.g. while walking through the community) and customer referrals, PAYG LPG customers may not be representative of the general population of the informal settlement.

However, a comparison of available SES characteristics of PAYG LPG households with that of a random sample from the same community revealed no significant differences in female occupation and household education levels (Table S3). Further, as individuals from the community in Eldoret had a higher SES than PayGo Energy customers living in the informal settlement in Nairobi, a 75% decline in days per month using full cylinder LPG in Eldoret signals that household SES likely does not explain differences in LPG usage patterns that occurred between PAYG LPG and full cylinder LPG users in Mukuru kwa Reuben during lockdown.

As per capita LPG consumption varies according to the level of urbanicity,^41^ PAYG LPG usage in an urban informal settlement may not manifest in peri-urban/rural communities due to differences in availability of LPG refills (e.g. frequency of LPG cylinder deliveries), baseline familiarity with LPG among residents and sociocultural barriers.^31,50^ Moreover, home delivery of LPG refills in peri-urban and rural communities with lower population density may lead to higher fuel surcharges transferred to customers due to longer travel distances and more complex distribution logistics, influencing customer retention. Issues related to infrastructure should be considered, as power outages/ connectivity issues with mobile service providers may be a significant barrier in rural communities, which can hamper customers’ ability to use mobile money for LPG payments. Improved cellular infrastructure can help alleviate reliability concerns.

This study did not include usage patterns among secondary cooking fuels. If stove stacking persists, levels of household air pollution likely remain above WHO guidelines.^12,51^ Household air pollution could increase above baseline levels during COVID-19 lockdown if households cook with polluting fuels for longer time periods, as identified from the stove monitoring data in Eldoret (Figure 5a). Fuel stacking can impact the profitability of commercial PAYG LPG companies, as their business models are dependent on consistent fuel consumption; low PAYG LPG usage will hinder revenue flows and can disincentivize future market entry. More research on stove stacking in the context of PAYG LPG is warranted.

## 5. Conclusion

PAYG LPG, a ‘leapfrog’ technology offering flexibility for cooking fuel purchase, was robust to consumers’ financial hardships induced by the COVID-19 control response by allowing them to adjust their payment patterns and consumption to meet changes in household income and family dynamics. PAYG LPG smart meter technology was also valued by customers for its safety, convenience, ease of payment and speed of cooking. These features make PAYG LPG a promising technology for promoting sustained use of clean cooking in communities where individuals work in the informal sector or experience seasonal income fluctuations. PAYG LPG should be actively considered in low and middle-income countries as one of the mechanisms for addressing the negative health and environmental impacts associated with reliance on polluting fuel for cooking.

## Data Availability

Data used in this study is not publicly available.

## Acknowledgements

The authors would like to acknowledge Ashley Berman from PayGo Energy for offering input on PayGo Energy’s customer recruitment process and providing a figure that depicts the company’s market entry timeline. The authors also acknowledge Joan Chepngeno, Mary Kiano, Sharon Cherono and Ruth Kemboi for helping translate the customer interviews from Kiswahili to English.

