## Supplemental Information for "Pay-as-you-go LPG supports sustainable clean cooking in Kenyan informal urban settlement, including during a period of COVID-19 lockdown"

### *PayGo Energy Path to Market*

PayGo Energy piloted their first version of liquefied petroleum gas (LPG) metered technology and began recruiting customers in 2016 (Figure S1). All onboarded PayGo Energy customers began to be supplied with pay-as-you-go (PAYG) LPG metered technology (called "Goldcard meters") in August 2017. In 2018, all PayGo Energy customers were using metered technology. In 2019, PayGo Energy began replacing the Goldcard meters with cylinder smart meters.

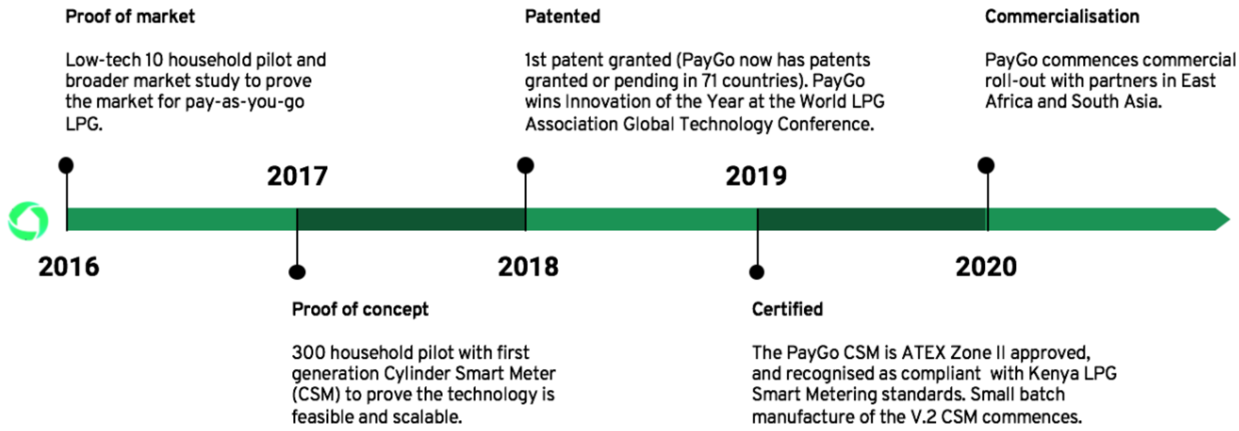


**Figure S1. Timeline of PayGo Energy's market entry**

### *Comparison of Smart Meter Technology used by PayGo Energy*

A sensitivity analysis (chi-square tests) was conducted with GoldCard metered data excluded to assess any changes to reported consumption statistics (Table S1). While monthly per capita PAYG LPG consumption among all PayGo Energy customers before lockdown was similar with that from customers only using cylinder smart meters (Goldcard meters excluded), PAYG LPG consumption per single cooking event was much lower with Goldcard metered data excluded (Table S1). This difference is due to rounding error as LPG usage data from Goldcard meters are less accurate (nearest 0.2 kilograms) than data from cylinder smart meters (nearest 0.01 kilograms). As households are more frequently consuming less gas than the 0.2 kg threshold for the Goldcard meters during a single cooking event (Table S1), Goldcard meters artificially inflated LPG consumption per cooking event. Mean monthly per capita PAYG LPG consumption was very similar between all PayGo Energy customers and only those using cylinder smart meters both before and during COVID-19 community lockdown.

**Table S1. Comparison of Pay-As-You-Go (PAYG) LPG fuel usage data using data from all customers (Goldcard Meters + Cylinder Smart Meters) (N=437) and only customers using Cylinder Smart Meters (N=239) before (January 2018-February 2020) and during (March 2020-June 2020) the COVID-19 lockdown**

| | (Mean (SD)) | | | | | |
|---|---|---|---|---|---|---|
| | Pre-Lockdown | | Pre-Lockdown | | Lockdown | |
| Metric | All Customer Data (N=437)[1] | Data From Smart Meters only (N=239) | All Customer Data (N=298)[2] | Data From Smart Meters only (N=189) | All Customer Data (N=298)[2] | Data From Smart Meters only (N=189) |
| Kg of gas used per cooking event | 0.17 (0.14) | 0.04 (0.05) | 0.15 (0.12) | 0.04 (0.05) | 0.11 (0.12) | 0.04 (0.07) |
| Kg of gas/capita/month | 0.97 (0.74) | 0.90 (1.0) | 0.98 (0.30) | 0.98 (0.28) | 1.25 (1.01) | 1.24 (0.47) |

1. All households that used PAYG LPG since January 2018
2. Only households that still had an active PAYG LPG account during the COVID-19 lockdown

There were not significant socioeconomic differences among all PayGo Energy customers and only customers upgraded to cylinder smart meters during the analysis period (Table S2). Thus, consumption of PAYG LPG during a single cooking event from only customers upgraded to cylinder smart meters is likely representative of the cooking event LPG consumption among all PayGo Energy customers.

**Table S2. Comparison of Socioeconomic Characteristics from all customers (Goldcard Meters + Cylinder Smart Meters) (N=437) and only customers using Cylinder Smart Meters (N=239) before (January 2018-February 2020) and during (March 2020-June 2020) the COVID-19 lockdown**

| Characteristic | All PayGo Energy Customers (n=437) | Customers using Cylinder Smart Meters (n=211) | Test statistic ($\chi^2$); P-value |
|---|---|---|---|
| **Sex of main cook** | | | 0.73 p=0.39 |
| Female | 211 (83%) | 48 (88%) | |
| **Sex of fuel decision-maker** | | 19 | 0.58 p=0.45 |
| Female | 153 (59%) | 36 (65%) | |
| **Highest Household Level of Education** | | | 2.29 p=0.32 |
| None | 4 (2%) | 1 (2%) | |
| Primary | 57 (26%) | 10 (22%) | |
| Secondary or university | 160 (72%) | 35 (76%) | |
| **Occupation (male head of household)** | | | 4.26 p=0.24 |

| | | | |
|---|---|---|---|
| Business/government employee | 22 (27%) | 6 (60%) | |
| Informal sector | 22 (27%) | 1 (10%) | |
| Day laborer/casual job | 17 (21%) | 2 (20%) | |
| Unemployed | 2 (2%) | 1 (10%) | |
| **Occupation (female head of household)** | | | 1.93<br>p=0.75 |
| Day laborer/casual job | 22 (26%) | 2 (13%) | |
| Business/government employee | 11 (13%) | 3 (20%) | |
| Informal sector | 39 (46%) | 9 (60%) | |
| Unemployed | 9 (11%) | 1 (6%) | |
| Farmer/homemaker | 3 (4%) | 0 | |

Note: some demographic data was only collected from a subset of participants. Totals for certain variables do not sum up to overall sample size.

#### *Socioeconomic Characteristics of PayGo Energy Customers*

Another sensitivity analysis (chi-square tests) was conducted to compare the socioeconomic status (SES) of PayGo Energy customers with that of a random sample of households participating in a previous study in the same community.[1] There were no significant differences found in highest household education level and the occupation of the female and male head of household (Table S3). However, a higher proportion of cooking fuel decision makers were female among PAYG LPG customers (59%) than those that were not (47%) (p=0.05) (Table S3). Further, the male head of household was more likely to be employed among PayGo Energy customers (98%) compared to the proportion of employed males in the community (88%). Additionally, the percent of male primary cooks using PAYG LPG (17%) was significantly higher than the percent of male cooks in the community not using PAYG LPG (8%).

**Table S3. Comparison of Pay-As-You-Go (PAYG) LPG users and non-PAYG LPG users in Mukuru Kwa Reuben informal settlement (Nairobi, Kenya)**

| Characteristic | PAYG LPG Users (n=435) | Non PAYG LPG Users (n=191) | Test statistic ($\chi^2$); P-value |
|---|---|---|---|
| **Sex of respondent** | | | 0.11<br>p=0.73 |
| Female | 308 (71%) | 130 (73%) | |
| **Sex of main cook** | | | 5.8<br>p=0.02* |
| Female | 211 (83%) | 163 (92%) | |
| **Sex of fuel decision-maker** | | | 4.0<br>p=0.05* |
| Female | 153 (59%) | 49 (47%) | |
| **Highest Household Level of Education** | | | 3.2<br>p=0.20 |
| None | 4 (2%) | 0 | |
| Primary | 57 (26%) | 51 (27%) | |
| Secondary or university | 160 (72%) | 140 (73%) | |
| **Occupation (male head of household)** | | | 10.8<br>p=0.01* |
| Business/government employee | 22 (27%) | 41 (33%) | |
| Informal sector | 22 (27%) | 21 (17%) | |
| Day laborer/casual job | 17 (21%) | 46 (38%) | |

| | | | |
|---|---|---|---|
| Unemployed | 2 (2%) | 15 (12%) | |
| **Occupation (female head of household)** | | | 3.2<br>p=0.52 |
| Day laborer/casual job | 22 (26%) | 19 (36%) | |
| Business/government employee | 11 (13%) | 8 (15%) | |
| Business owner | 39 (46%) | 17 (33%) | |
| Unemployed | 9 (11%) | 7 (13%) | |
| Farmer/homemaker | 3 (4%) | 1 (2%) | |
| **Household Size (number of rooms)** | | | 74.4<br>p=<0.001* |
| 1 | 124 (49%) | 156 (88%) | |
| 2 | 68 (27%) | 18 (10%) | |
| 3+ | 59 (24%) | 4 (2%) | |
| **Main lighting fuel** | | | |
| Electricity | 169 (73%) | 165 (93%) | 31.5<br>p=<0.001* |
| Kerosene, candles, other | 73 (27%) | 13 (7%) | |

*=statistically significant at alpha=0.05 level
Note: some demographic data was only collected from a subset of participants. Total for certain variables do not sum up to overall sample size.

An additional sensitivity analysis was conducted to compare PAYG LPG customers in Mukuru kwa Reuben with LPG users in Eldoret, Kenya to determine if higher use of LPG during lockdown may be related to socioeconomic differences between the two populations. The results indicated a significantly higher SES among households in Eldoret compared with Mukuru kwa Reuben across all relevant variables examined (education, occupation, household size, electricity access for lighting) (Table S4). Thus, higher maintenance of LPG usage during lockdown among the lower SES households in Mukuru kwa Reuben is likely the result of the payment scheme offered by PAYG and not due to higher income. As households in Eldoret dramatically reduced their LPG consumption during lockdown, PAYG LPG may be beneficial in preventing increases in polluting fuel use among higher income households. It is evident that polluting cooking fuel use likely increased across various income strata in Kenya, warranting the need for technological and policy solutions that can be scaled up to large sectors of the population.

**Table S4. Comparison of Pay-As-You-Go (PAYG) LPG users in Mukuru Kwa Reuben informal settlement (Nairobi, Kenya) (n=435) and LPG users in Eldoret, Kenya (n=201)**

| **Characteristic** | **PAYG LPG Users (n=435)** | **LPG Users in Eldoret, Kenya (n=201)** | **Test statistic ($\chi^2$); P-value** |
|---|---|---|---|
| **Highest Household Level of Education** | | | 38.2<br>p<0.001* |
| None | 4 (2%) | 7 (3%) | |
| Primary | 57 (26%) | 41 (20%) | |
| Secondary or university | 160 (72%) | 153 (76%) | |
| **Occupation (male head of household)** | | | 20.4<br>p<0.001* |
| Business/government employee | 22 (27%) | 81 (51%) | |
| Business owner/Informal sector | 22 (27%) | 50 (31%) | |

| | | | |
|---|---|---|---|
| Day laborer/casual job | 17 (21%) | 24 (15%) | |
| Unemployed | 2 (2%) | 4 (3%) | |
| **Occupation (female head of household)** | | | 29.8<br>P<0.001* |
| Day laborer/casual job | 22 (26%) | 1 (2%) | |
| Business/government employee | 11 (13%) | 15 (35%) | |
| Business owner/Informal sector | 39 (46%) | 18 (42%) | |
| Unemployed | 9 (11%) | 4 (9%) | |
| Farmer/homemaker | 3 (4%) | 5 (12%) | |
| **Household Size (number of rooms)** | | | 41.4<br>p<0.001* |
| 1 | 124 (49%) | 38 (19%) | |
| 2 | 68 (27%) | 54 (27%) | |
| 3+ | 59 (24%) | 109 (54%) | |
| **Main lighting fuel** | | | 32.4<br>p=<0.001* |
| Electricity | 169 (73%) | 185 (92%) | |
| Kerosene, candles, other | 73 (27%) | 16 (8%) | |

### *Recruitment of Customers*

The majority of PayGo Energy customers were recruited via door-to-door recruitment (n=156; 37%), PayGo Energy staff members identifying a new customer (e.g. on a walk through the community) (n=126; 30%) or referral by other customers (n=78; 19%) (Table S5).

**Table S5. PayGo Energy customer recruitment methods (N=417)**

| **Method** | **N (%)** |
|---|---|
| Door-to-door recruitment | 156 (37%) |
| PayGo Energy staff identified new customer | 126 (30%) |
| Customer referral | 78 (19%) |
| Marketing event | 28 (7%) |
| Customer initiated | 19 (5%) |
| Staff referral | 10 (2%) |

PayGo Energy customers included in the study sample were mostly recruited in two 'waves' in 2017 and 2019. This resulted in the majority of households having used PAYG LPG for approximately three years (31-36 months) or one year (7-12 months) (Table S6) by June 2020.

**Table S6. Months of smart meter data available for the study sample (N=386)**

| Number of months of PAYG LPG smart meter data available | Number of customers |
|---|---|
| 1-6 | 63 |
| 7-12 | 120 |
| 13-18 | 11 |

| | |
|---|---|
| 19-24 | 24 |
| 25-30 | 28 |
| 31-36 | 121 |
| 37-42 | 19 |

#### *Account Deactivations*

One in five customers who deactivated their PayGo Energy account cited high costs as their main reason; 13% (n=17) of respondents cited high costs specifically and 7% (n=9) switched to purchasing LPG under the cylinder recirculation model (CRM) in 2017, when the cost of PayGo LPG was higher than purchasing an LPG cylinder (Table S7). Account deactivations due to high costs suggests that price elasticity pays a key role in decision making of cooking fuel purchases. High price elasticity around use of modern cooking fuels has been documented in Brazil - around three million households using LPG for cooking switched to firewood or charcoal between 2016-2018 because of LPG price increases and reduced income due to higher unemployment.[2]

Among households using kerosene prior to registering for PAYG LPG, one-third (n=6) of those terminating their account with PayGo Energy due to high recurring fuel cost indicated switching to buying full LPG refills, while two-thirds (n=13) did not. This illustrates that introduction to LPG under a PAYG model did not consistently lead to sustained usage of LPG when prices were not satisfactory but was successful as entry point to start using LPG.

**Table S7. Reasons for PayGo Energy customers deactivating their account**

| **Reason** | **Overall** | **LPG** | **Kerosene** | **Charcoal** |
|---|---|---|---|---|
| Moved | 41 (33%) | 0 | 36 (35%) | 4 (67%) |
| Tampered or stolen equipment | 34 (27%) | 3 (27%) | 29 (28%) | 0 |
| Switched to standard LPG refills under the BCRM[1] | 9 (7%) | 3 (27%) | 6 (6%) | 0 |
| Too expensive | 17 (13%) | 3 (27%) | 13 (13%) | 0 |
| Stopped using gas (reason unknown) | 24 (19%) | 2 (18%) | 19 (18%) | 2 (33%) |

1.BCRM = branded cylinder recirculation model. This refers to buying and then swapping a full LPG cylinder once empty with a full filled cylinder of the same size and brand.
Note: reasons for deactivation across primary fuel types do not always add up to overall category due to missing data on previous primary fuel from some customers

#### *Sex of Fuel Decision Maker*

Similar to an higher proportion of female primary cooking fuel decision-makers among households adopting PAYG LPG compared with the population-level proportion of female primary cooking fuel decision-makers in the community (Table S3), a significantly higher proportion (76%) of cooking fuel decision makers were female among households previously bulk purchasing LPG prior to registering with PayGo Energy, compared with those that did not (55%) (Table S8).

**Table S8. Sex of fuel decision-maker by households using or non-using LPG prior to registering with PayGo Energy**

| | Non-LPG user prior to registering for PAYG LPG (N=206) | LPG user prior to registering for PAYG LPG (N=29) | P-value ($\chi^2$) |
|---|---|---|---|
| **Sex of fuel decision-maker** | | | |
| **Female** | 113 (55%) | 22 (76%) | 0.05* |
| **Male** | 93 (45%) | 7 (24%) | |

*=statistically significant at alpha=0.05 level
Note: information on sex of the cooking fuel decision-maker was only collected from a subset of participants.

### *Cooking Event Patterns*

Among 20 households that previously used kerosene and LPG under the CRM before adopting PAYG LPG, there was a notable increase in the number of cooking events per day from once to twice a day (Figure S2).

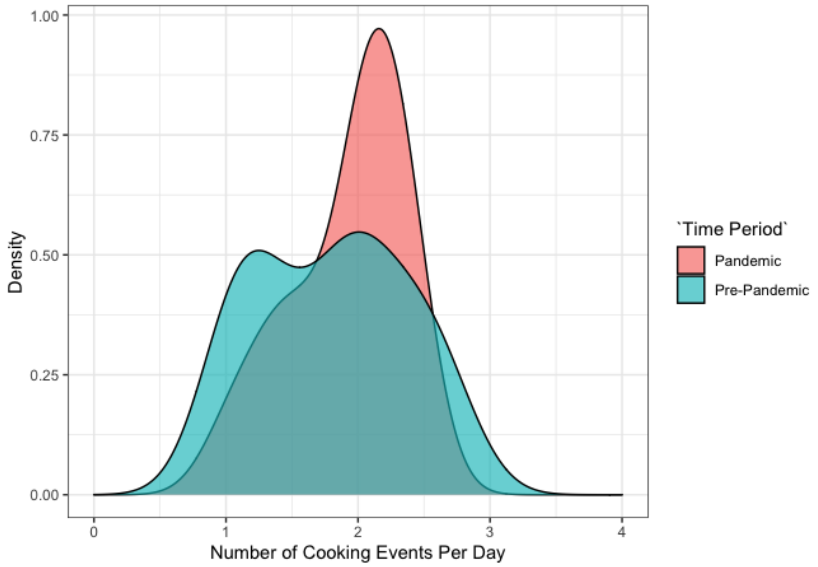


**Figure S2. Number of cooking events per day using PAYG LPG stove before ('Pre-Lockdown) and during COVID-19 community lockdown ('Lockdown') among households previously using LPG and kerosene stoves before registering with PayGo Energy (n=20).**

#### *Hour of Day Cooking with Pay-As-You-Go LPG*

The hour of day when PAYG LPG was used varied according to the previous primary cooking fuel used before adopting PAYG LPG. PAYG LPG was commonly used for preparing breakfast (6-8 am) and lunch (1-2 pm) among households using charcoal before registering with PayGo Energy. PAYG LPG was used more commonly for preparing dinner (6-8 pm) by households previously cooking with kerosene or LPG (Figure S3).

PAYG LPG was most commonly used for preparing breakfast (6-9 am) and dinner (7-10 pm) among households where the female head was employed in the informal sector (Figure S4). Among those formally employed, PAYG LPG was more commonly used for preparing breakfast before the lockdown (6-8 am) with a shift to cooking with PAYG LPG more commonly for lunch (12-2 pm) and dinner (6-8 pm) before the lockdown. This likely reflects individuals eating their lunch at home during lockdown when they would regularly eat this meal at work pre-lockdown. PAYG LPG cooking times at breakfast, lunch and dinner were less common among those with casual jobs, perhaps due to less structured work schedules (Figure S4).

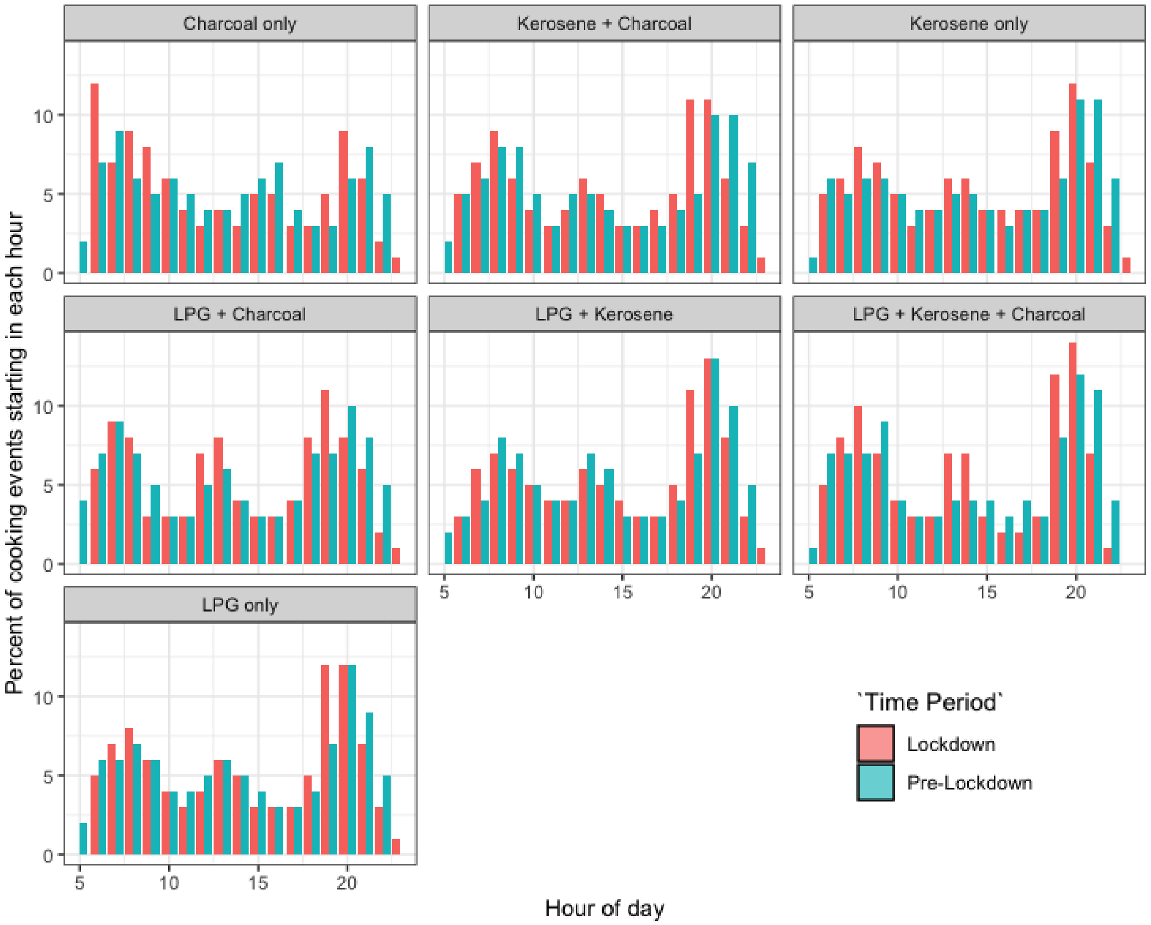


**Figure S3. Hour of day of cooking events registered to PAYG LPG smart meter technology by previous primary cooking fuel used when registering with PayGo Energy before ('Pre-Lockdown) and during COVID-19 community lockdown ('Lockdown')**

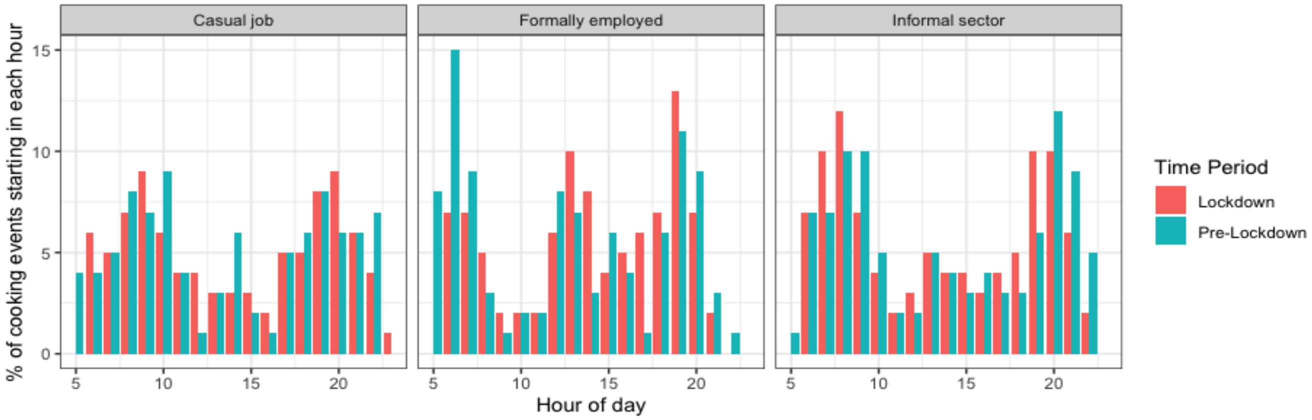


**Figure S4. Hour of day of cooking events registered to PAYG LPG smart meter technology by female occupation when registering with PayGo Energy before ('Pre-Lockdown) and during COVID-19 community lockdown ('Lockdown')**

### *Cooking and Spending Patterns among Commercial Customers*

Monthly consumption of PAYG LPG among commercial customers decreased by 4 kg during lockdown (17.1 kg (pre-lockdown) to 13.1 kg (lockdown)) (Table S9). This contrasts with an increase in consumption of 0.6 kg per month among residential customers. The commercial cook interviewed stated that they decreased their PAYG LPG consumption due to a diminishing number of customers during this time period, particularly because of the dusk-to-dawn curfew, which prevented community members from dining at his restaurant at dinnertime.

Both residential and commercial customers decreased their average single payment amount; however, commercial customers decreased their spending by a wider margin. This resulted in commercial customers spending less on PAYG LPG per month during lockdown, while residential customers slightly increased their monthly expenditure.

**Table S9.** Pay-As-You-Go (PAYG) LPG fuel usage and spending habits among commercial and residential customers before (March-June 2018/2019) and during (March 2020-June 2020) the COVID-19 lockdown

| **Metric (Mean (SD))** | **Customer type** | | | |
|---|---|---|---|---|
| | **Commercial** | | **Residential** | |
| | **Pre-Lockdown** | **Lockdown** | **Pre-Lockdown** | **Lockdown** |
| Kg gas consumed per month | 17.1 (12.1) | 13.1 (10.7) | 3.2 (1.8) | 3.8 (2.2) |
| Cooking events per day | 2.06 (0.69) | 1.94 (0.58) | 1.07 (0.23) | 1.72 (0.65) |
| Minutes per cooking event | 15.4 (8.5) | 11.1 (4.2) | 14.4 (13.9) | 13.5 (13.0) |
| Single payment amount (KSh) | 218 (243) | 156 (202) | 336 (286) | 178 (189) |
| Amount spent per month (KSh) | 2534 (1649) | 2290 (1717) | 867 (469) | 816 (510) |
| Days between payments (median [IQR]) | 2.0 [0.9, 3.2] | 0.7 [0.3, 0.9] | 8.0 [4.0, 22.0]] | 4.0 [2.5, 9.0] |

#### *Cooking Intensity*

Cooking intensity (defined as grams of LPG used per minute) significantly increased during COVID-19 lockdown (Table 2). This may reflect differences in the type of meals cooked during the lockdown. There was not a significant correlation between cooking event length and cooking intensity (Figure S5).

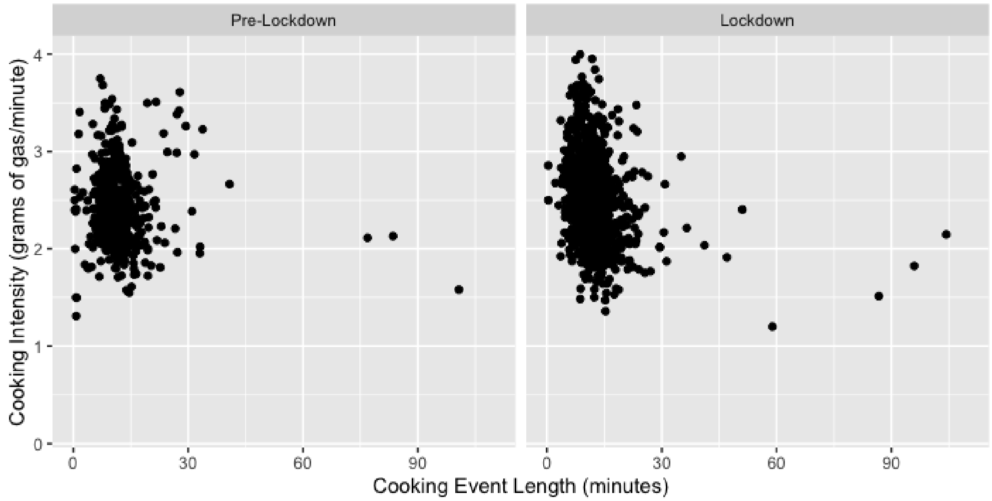

**Figure S5. Correlation of cooking eventh length and cooking intensity before ('Pre-Lockdown) and during COVID-19 community lockdown ('Lockdown')**

### *Intraclass Correlation*

There was higher within-household variability in the number of days using gas each month than between households (ICC <0.5) both before (0.41), and to a greater extent, during the lockdown (0.25) (Table S10). There was higher variability in payment amount between households than within households (ICC >0.7), showing that households appear to maintain a consist payment scheme when using PAYG LPG. The within-household variability in frequency of payments decreased substantially (higher ICC) during the COVID-19 lockdown (ICC=0.77) compared with before the lockdown (ICC=0.52).

**Table S10. Intraclass correlation coefficient (ICC) for PAYG LPG consumption and spending metrics**

| Characteristic | ICC[1] | | |
|---|---|---|---|
| | Pre-Lockdown | Pre-Lockdown (March-June 2018 & March-June 2019) | Lockdown (March-June 2020) |
| Kg/month/capita | 0.51 | 0.53 | 0.56 |
| Number of days using gas/month | 0.41 | 0.34 | 0.25 |
| Single payment amount | 0.78 | 0.82 | 0.73 |
| Number of payments/month | 0.63 | 0.52 | 0.77 |

1.ICC represents the variability between households relative to the total variance between and within households.
